# Assessing the Utility of Low Resolution Brain Imaging: Treatment of Infant Hydrocephalus

**DOI:** 10.1101/2021.07.21.21260949

**Authors:** Joshua R. Harper, Venkateswararao Cherukuri, Tom O’Reilly, Mingzhao Yu, Edith Mbabazi-Kabachelor, Ronald Mulando, Kevin N. Sheth, Andrew G. Webb, Benjamin C. Warf, Abhaya V. Kulkarni, Vishal Monga, Steven J. Schiff

## Abstract

As low-field MRI technology is being disseminated into clinical settings around the world, it is important to assess the image quality required to properly diagnose and treat a given disease and evaluate the role of machine learning algorithms, such as deep learning, in the enhancement of lower quality images. In this post-hoc analysis of an ongoing randomized clinical trial, we assessed the diagnostic utility of reduced-quality and deep learning enhanced images for hydrocephalus treatment planning. CT images of post-infectious infant hydrocephalus were degraded in terms of spatial resolution, noise, and contrast between brain and CSF and enhanced using deep learning algorithms. Both degraded and enhanced images were presented to three experienced pediatric neurosurgeons accustomed to working in low- to middle-income countries (LMIC) for assessment of clinical utility in treatment planning for hydrocephalus. In addition, enhanced images were presented alongside their ground-truth CT counterparts in order to assess whether reconstruction errors caused by the deep learning enhancement routine were acceptable to the evaluators. Results indicate that image resolution and contrast-to-noise ratio between brain and CSF predict the likelihood of an image being characterized as useful for hydrocephalus treatment planning. Deep learning enhancement substantially increases contrast-to-noise ratio improving the apparent likelihood of the image being useful; however, deep learning enhancement introduces structural errors which create a substantial risk of misleading clinical interpretation. We find that images with lower quality than is customarily acceptable can be useful for hydrocephalus treatment planning. Moreover, low quality images may be preferable to images enhanced with deep learning, since they do not introduce the risk of misleading information which could misguide treatment decisions. These findings advocate for new standards in assessing acceptable image quality for clinical use.

## 1 Introduction

With an estimated 400,000 new cases worldwide each year, childhood hydrocephalus is the most common pediatric condition requiring neurosurgery globally [1]. Over 90% of cases occur in low- and middle-income countries (LMIC) [1]. In sub-Saharan Africa, approximately 180,000 infants per year are affected [2]. Hydrocephalus is characterized by a build up of intracranial cerebrospinal fluid (CSF) that, in infants, causes the head to enlarge. These infants need surgical treatment to survive requiring intracranial imaging for planning. In planning surgery it is important to know where the CSF is in relation to brain, and how many compartments are loculated where fluid is trapped. An imaging technology capable of showing contrast between brain and CSF at an appropriate resolution is required. We have previously suggested that a voxel size approaching 100 *mm*^3^ (e.g 3×3×10 mm3) could be sufficient for planning treatment [3].

The brain is an organ where the soft tissue and fluid encased within the skull have limited alternatives for imaging. Ultrasound is only effective within the first year of life before skull fusion closes the acoustical windows of the fontanels. The ionizing radiation associated with CT poses exceptional risks to infants [4, 5]; however, in sub-Saharan Africa CT is more prevalent than MRI [6] due to its lower cost. Although MRI is the gold standard for pediatric neuro-imaging, the high cost, strict siting requirements, and demanding maintenance schedule render high-field cryogenic systems infeasible for most of the developing world [6–9].

According to a 2014 baseline country survey on medical devices conducted by the World Health Organization, Uganda has 0.45 CT machines per million people and only 0.08 MRI machines per million people. By comparison, a high income country such as the Netherlands, has 12 CT and 12 MRI machines per million people (roughly 27 times more CT/million and 150 times more MRI/million people) [6]. Placed in the context of new hydrocephalus cases per year, with rates at least 10 times more per year in Africa than in Europe [1], the clinical need for globally sustainable diagnostic imaging devices is clear. Low-field MRI devices have been recently developed that are feasible for the developing world and show diagnostic promise for the treatment and management of illnesses such as hydrocephalus [3, 10–12].

The quality of an MRI image ultimately depends on the signal-to-noise ratio (SNR) per voxel. Higher field strength systems (>1.5 Tesla) can produce increased signal-to-noise pushing voxel size as low as hundreds of micrometers [13]. Low-field systems (<0.1 Tesla) inherently suffer from low signal-to-noise placing limits on achievable voxel size and including more baseline noise than most clinicians are accustomed to. Figure 1 demonstrates the difference in brain image quality between a high-field (Figure 1A) and a low-field (Figure 1B) MRI system.

**Figure 1:**
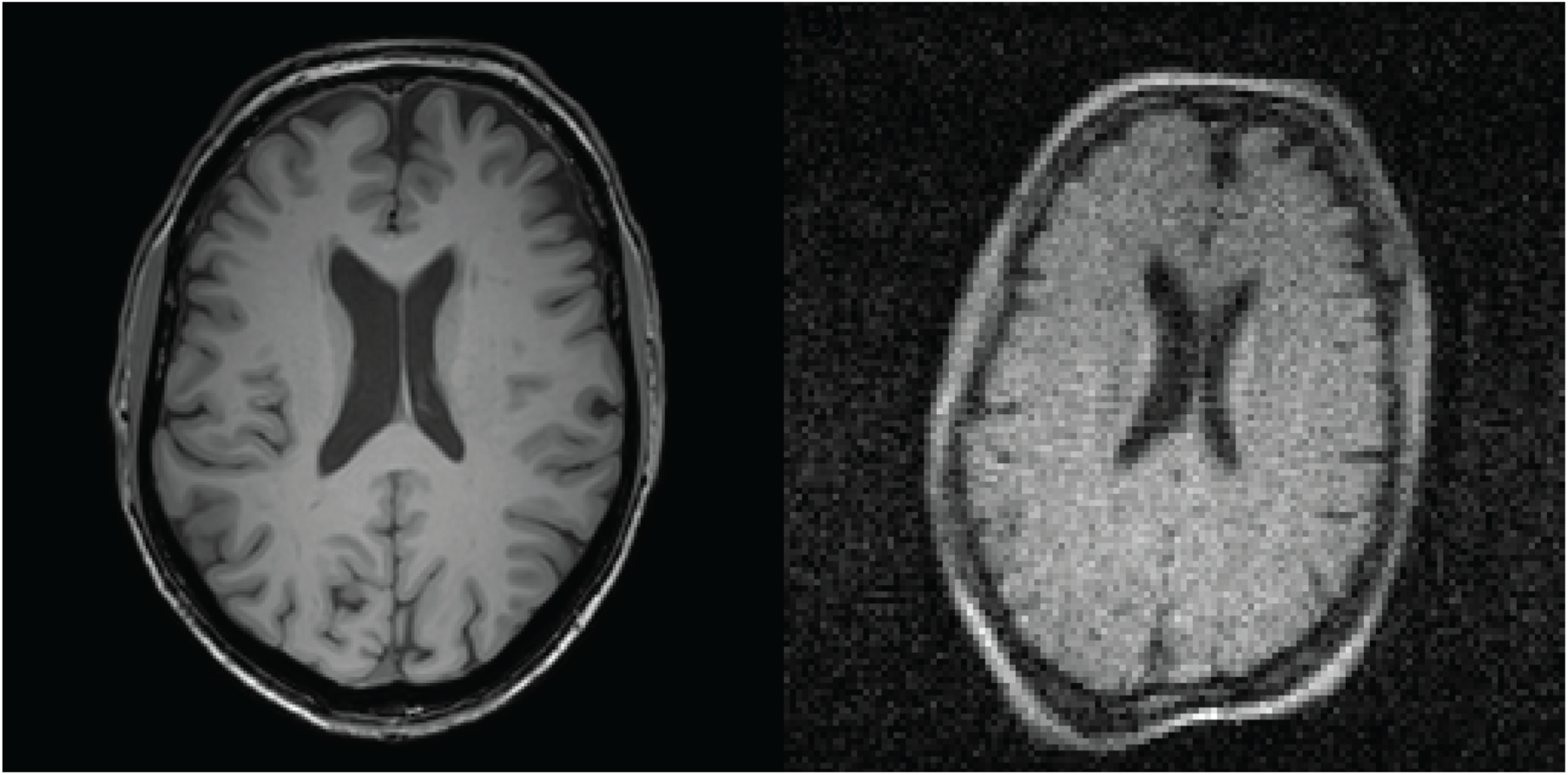
A comparison of the image quality between a high-field (3T) and a low-field (0.05 T) image of the brain of the same volunteer taken at the Leiden University Medical Center. A) A 256×256 3D T1 weighted TFE with Field of View: 200×175×156 mm, Resolution: 1.15×1.15×1.2 mm, TR/TE/TI = 9.8 ms/4.6ms/1050ms, ETL = 166, scan duration: 3 minutes 13 seconds; B) A 128×128 image at 0.05 T with Field of view: 256×256×200mm, Resolution: 2×2×4 mm, TR/TE = 400ms/15 ms, echo train length = 6, scan duration: 7 minutes 7 seconds.

The adoption of low-field MRI into clinical practice depends largely on a longstanding and recently growing body of evidence that higher image quality does not always lead to better diagnostic accuracy or better patient outcome [14]. In clinical practice there exists a threshold of image quality for specific pathologies, above which no further outcome-based value can be observed [15]. It has been demonstrated that 0.5 Tesla MRI can be as diagnostically accurate as 1.5 Tesla MRI for a variety of diseases including central nervous system pathologies [16], heptic lesions [17], and multiple-sclerosis [18]. It has also been shown that a 0.064 Tesla MRI can have comparable diagnostic accuracy to a 1.5 Tesla MRI for neoplasms and white matter disease [19]. Although the threshold of image quality required to plan effective hydrocephalus treatment has not been previously explored, we hypothesized that the level of resolution, tissue contrast, and SNR provided by CT or high-field MRI substantially exceeds this threshold.

Various machine learning-based methods have previously been used to perform super-resolution enhancement of low-quality MRI images. Interpolation based methods [20] are simple to implement but lack prior information often resulting in blurring. Model-based methods [21–23] explore the stochastic mechanism in the MRI generating process and model it with prior information; nevertheless, the design of a suitable regularization for the model can be difficult. Learning-based methods have the advantage of modeling and learning the mapping of low-quality images to high-quality images from data alone [24–27]. Recently, deep learning has shown impressive performance in the field of super-resolution of MRI [28–32].

In the present work, we assess the diagnostic utility of reduced-quality and deep learning enhanced images for hydrocephalus treatment planning. We focus on the most common form of infant hydrocephalus in sub-Saharan Africa – postinfectious [33]. This form of hydrocephalus is uncommon outside of LMIC [1], and the only abundant high-resolution comparative images are from CT. We developed an image utility assessment which was completed by three senior neurosurgeons with extensive experience in the treatment and management of hydrocephalus in low-resource settings [33–35]. Qualitative and quantitative measures of image utility are used to classify images revealing the quality threshold for treatment planning of hydrocephalus in terms of resolution, noise, and contrast between brain and CSF. We further evaluate how machine learning can lead to misleading modifications during the enhancement of low-resolution imagery.

## 2 Methods

Three experienced pediatric neurosurgeons accustomed to working in LMIC, with particular experience in interpretation of postinfectious hydrocephalus imagery of African infants, were chosen as participants in the image utility assessment. CT images were acquired from a repository of 90 patients enrolled in an ongoing randomized clinical trial (median age of 3.1 months, 39% female [34]) and treated at the CURE Children’s Hospital of Uganda for post-infectious hydrocephalus. The center-most image slice from each patient was chosen for the assessment as either a test image (10 randomly selected, Figure S5) or a learning library image (remaining 80). The images are 512×512 with 0.4 mm resolution (20.48 cm field of view). Each slice is 5 mm thick.

The 10 test images were degraded in terms of resolution, noise, and contrast between brain and CSF. Since the field of view remained constant for all images, resolution was adjusted by reducing the matrix size of the image. Because of this relationship, we adopt the term “resolution” to describe changes in image matrix size for the present work. An image parameter space, as shown in Figure 2A-B, was constructed consisting of the variables: 1) resolution (32×32, 64×64, 128×128, 512×512); 2) contrast reduction (20 levels between 0 and 1), 3) and noise added (20 levels between 0 and 1) resulting in 1,600 possible parameter combinations.

**Figure 2:**
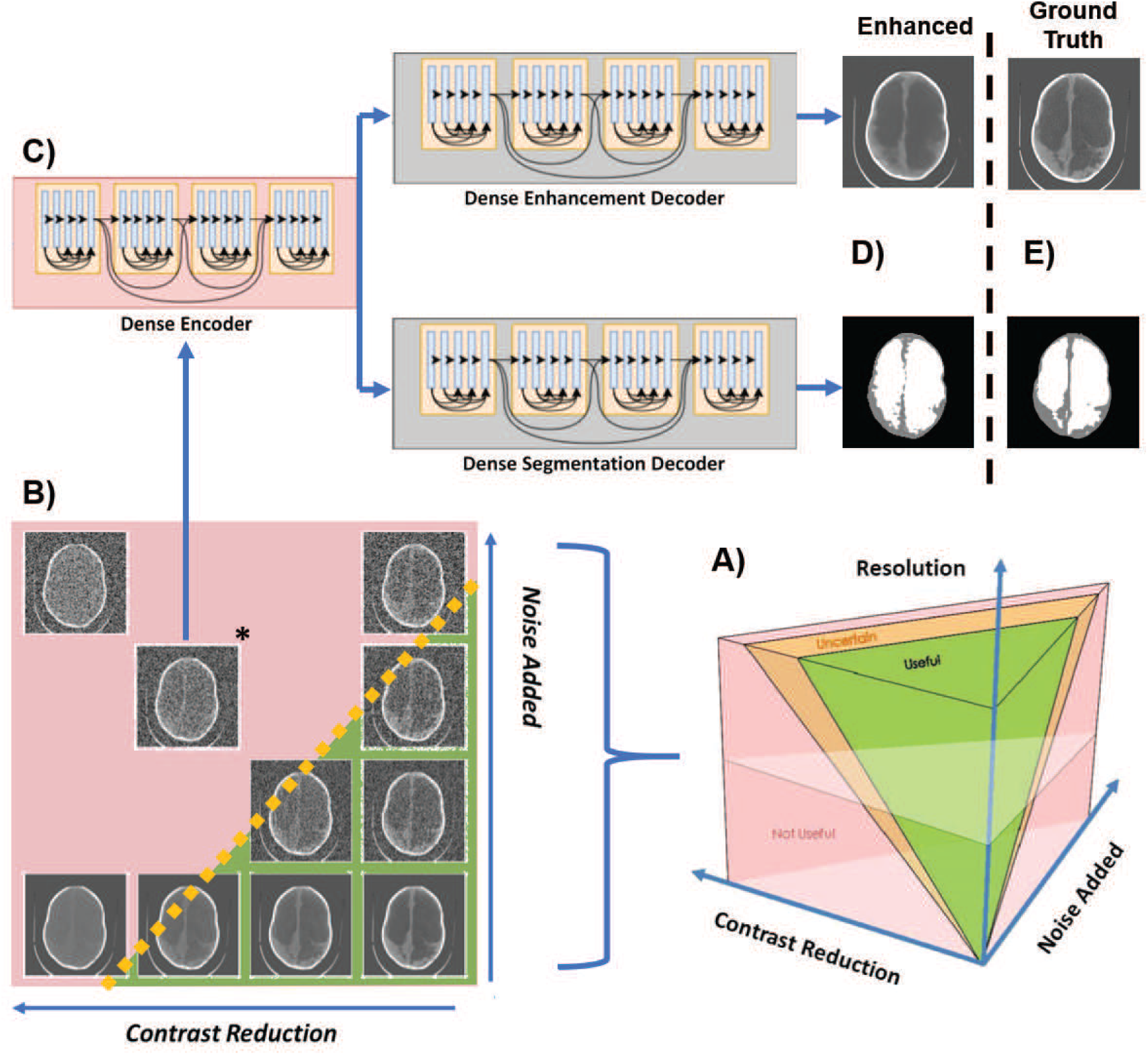
Schematic of study. In A) the image parameter space describing all possible combinations of noise, contrast between brain and CSF, and image resolution are visualized. There is likely to be a region of parameter combinations yielding images which are useful for hydrocephalus treatment planning (green volume), a region of parameter combinations that are not useful (red volume), and region of uncertainty in between (orange volume). In B) we show a single plane from image parameter space in which all images have 512×512 resolution. The lower right corner has maximum contrast between brain and CSF and least noise considered in this study and the upper left corner has the lowest contrast and most noise. In C) the starred image from panel B) is chosen to be enhanced with a single encoder dual decoder (SEDD) architecture following the DenseNet network described in [31, 36]. The output of such enhancement is seen in the upper panel of D) with corresponding segmentation in the lower panel of D). The ground truth version of the enhancement and segmentation from the original image without degradation or enhancement is shown in E) and called “ground truth”.

Resolution was down-sampled from the 512×512 image using bi-linear interpolation. The averaging between pixels in bi-linear interpolation can be considered an approximation of a partial volume effect.

Contrast between brain and CSF was reduced using histogram compression (Figure S6), an algorithm developed specifically for this purpose. In histogram compression the histogram of gray-scale values for brain and CSF are iteratively compressed into a smaller gray-scale bandwidth to simulate loss in tissue contrast.

Gaussian noise with mean equal to variance was added according to known noise characteristics of CT images [37]. Since lower resolution images are more sensitive to noise, the noise added was scaled by clinical inspection for each resolution so that both useful and not useful images would be represented. The noise variance added was scaled by resolution as follows and normalized to the maximum value: from 0 to 0.001 (32×32), 0 to 0.01 (64×64), 0 to 0.05 (128×128, and 0 to 0.13 (512×512).

In [31, 32], deep learning networks took advantage of low-rank structural prior information to enhance low quality images. Building on this work, we developed a deep learning network capable of simultaneously enhancing and segmenting CT images of infant hydrocephalus that have been artificially degraded. Following the DenseNet network described in [38], a single encoder dual decoder (SEDD) architecture was used to enhance CT images that have reduced quality. Deep learning networks, as shown in Figure 2C-E, were trained for two resolutions (64×64 and 128×128) at seven locations in parameter space using library images. With noise added as the x-coordinate and contrast reduction as the y-coordinate, networks were trained for both resolutions at: 1) (0.3,0.3), 2) (0.6,0.3), 3) (0.3,0.6), 4) (0.6,0.6), 5) (0.9,0.6), 6) (0.6,0.9), 7) (0.9,0.9) (Figure S7). The least degraded network is network 1. The networks were built by degrading the 80 library images at each of the 14 network locations and training with the original non-degraded image as ground truth. After training, the 10 test images were degraded at the network locations and enhanced generating 140 deep learning enhanced images.

From the 1,600 parameter combinations applied to the 10 test images, 420 cases were randomly presented to the panel of experts along with all 140 deep learning enhanced images. The image utility assessment was divided into two parts. In Part 1, the images were shown in 140 panels of 4 images each, as shown in Figure 3A. In each of the 140 panels, one image location was randomly selected for an enhanced image and the other three were degraded images. The expert was not told that there would be enhanced images. In each panel, the expert was asked to select which, if any, of the 4 images are clinically useful for planning hydrocephalus treatment (see Supplementary Methods for full instructions). The data from the three experts were combined by addition of scores for each image in order to be classified as useful, uncertain, or not useful. If all three experts agreed that an image was useful, this image received a 3 (i.e. Useful). If all experts agreed that an image was not useful this image received a 0 (i.e. Not Useful). Uncertain images received a score of either 1 or 2.

**Figure 3:**
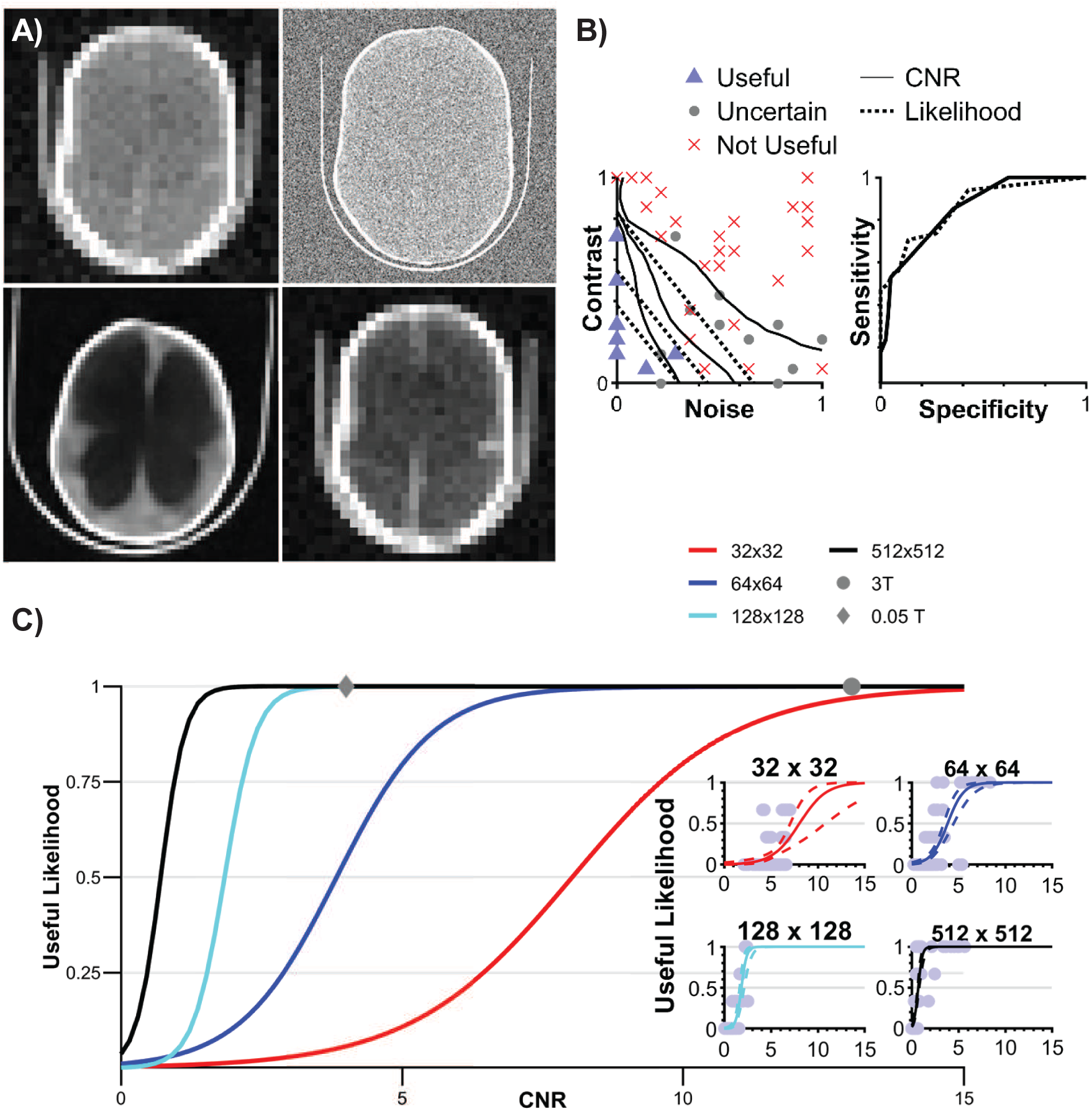
The figure shows results from Part 1 of the Assessment. In A) we show an example panel from Part 1 of the assessment. The lower left image is an enhanced image and all other images are degraded. The experts must indicate which (if any) is useful. The left panel of B) shows raw classification data from Part 1 for 64×64 images. Solid lines are lines of constant contrast-to-noise ratio (CNR). Dashed lines show lines of constant usefulness likelihood from the multivariate logistic regression. The right panel of B) shows the receiver operating characteristic curves. In C) we show the univariate logistic regression models for each resolution with CNR as the predictor. The diamond and circle datapoints show the calculated CNR values for the low-field and high-field MRI images shown in Figure 1, respectively. Resolution for these images lie between the 128×128 and 512×512 curves, which overlap for the CNR values reported. The bottom four panels of C) show the raw classification data for each resolution.

In Part 2, the experts were shown enhanced images in a side-by-side comparison with their corresponding 512×512 non-degraded versions as seen in Figure 4A. The experts were asked to assess whether the spatial errors in the enhanced version were acceptable or would alter treatment decisions (see Supplementary Methods for full instructions). The data from Part 2 were also combined by addition of scores. Part 2 enhanced images receiving a 3 were classified as useful (i.e. useful in both Part1 and Part2), those receiving a 1 or 2 were classified as uncertain, and those receiving a 0 were classified as misleading (i.e. useful in Part 1, but shown to have unacceptable error in Part 2).

**Figure 4:**
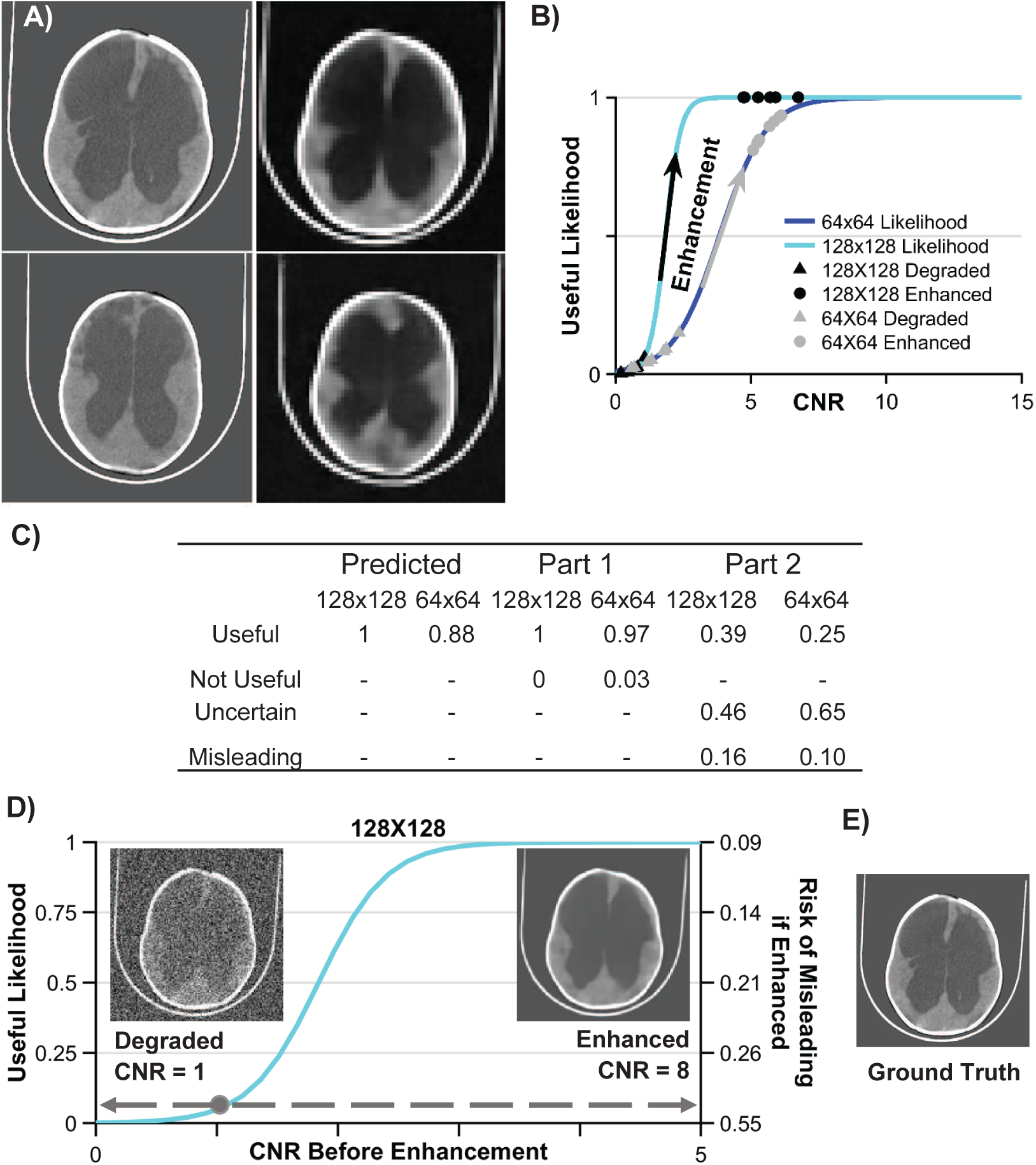
The figure shows results from Part 2 of the assessment. A) An example panel from Part 2 of the assessment. The left column of images are ground truth and the right column are the enhanced versions. B) shows the usefulness likelihood curves based on image CNR. The triangles show the average CNR for each network location before enhancement and the circles show the average CNR for each network after enhancement. C) shows the predicted usefulness likelihood of the enhanced images based on CNR after enhancement, the actual Part 1 classification of the enhanced images, and the Part 2 re-classification of the enhanced images after comparison with ground truth. In D) we compare the usefulness likelihood of the degraded images with the risk of a misleading result if the image is enhanced for 128×128 images. The left vertical axis shows the usefulness likelihood of the degraded image and the right vertical axis shows the risk of a misleading result if the corresponding degraded image were enhanced. In D) we also show an example degraded image on the left with CNR = 1, the enhanced version of this image on the right with CNR = 8 after enhancement and corresponding high likelihood of misleading results after enhancement. Finally, E) shows the ground truth version of the example image in D) for comparison.

In addition, an analysis of inter-rater reliability was performed using a variation of Cohen’s Kappa, as described in [39], which accounts for the existence of prevalence in the data and bias between evaluators (see Supplementary Methods). For this analysis, the data was divided into three parts: 1) classification of Part1 degraded images, 2) classification of Part 1 enhanced images, and 3) classification of Part 2 enhanced images. The Kappa statistic of [39] was calculated for all possible pairings of evaluators and conclusions regarding agreement were drawn based on the interpretation of Kappa values as suggested in [39] and [40].

Univariate and multivariate logistic regression was used to investigate the ability of contrast, noise, and contrast-to-noise ratio to predict image classification. A deviance statistic was used to assess goodness of fit of the logistic regression models. The deviance of the model is a chi-squared statistic which assesses the difference between the maximum log likelihood of the chosen model and that of the null model (i.e. the average probability of a classification at a given resolution being useful).

## 3 Results

### 3.1 Part 1: What makes an image useful?

We first characterize the relationship between resolution, contrast, noise and usefulness. The inter-rater reliability for the classification of degraded images shows fair agreement between evaluators 1 and 2 (K = 0.33), substantial agreement between evaluators 2 and 3 (K = 0.94), and fair agreement between evaluators 1 and 3 (K = 0.36). For all three evaluators, there was a high prevalence for classifying Part 1 enhanced images as being useful (see Supplementary results). As such, inter-rater reliability calculations for this data are not informative and all evaluators are in near perfect agreement. In Figure 3A we show several degraded images, of which the lower left is enhanced by deep learning. The left panel of Figure 3B shows how the contrast and noise of each image relates to the image classification determinations at 64×64 resolution (see Figure S10 for full dataset results). The solid contour lines in Figure 3B show lines of constant contrast-to-noise ratio between brain and CSF averaged from the full dataset of images. In comparison, the dotted lines show constant usefulness likelihood based upon a multivariate logistic regression model with contrast and noise as predictors. For images with each of the four resolutions considered, the multivariate logistic regression model provided a significant fit with p-values less than 0.01 (p_32×32_ = 7e-6, p_64×64_ = 4e-27, p_128×128_ = 2e-17, p_512×512_ = 8e-32). Note that there is qualitative agreement between the average contrast-to-noise contours and the lines of constant likelihood that the image is useful. On the right of Figure 3B receiver operating characteristic curves demonstrate that average contrast-to-noise and likelihood are both comparably effective classifiers of image utility with areas under their curves > 0.85 (curves for full dataset in Figure S11).

Since average contrast-to-noise appeared to be an effective classifier, Figure 3C shows that individual image contrast-to-noise alone is a significant predictor of usefullness likelihood, stratified by resolution. The grey circle shows the usefullness likelihood of the 256×256 brain image from the 3 Tesla system in Figure 1A based on its contrast-to-noise ratio (CNR=13). The grey diamond shows the same for the 128×128 brain image from the 0.05 Tesla system in Figure 1B (CNR=4). Though the image generated by the 3T system has twice the resolution and 3 times the CNR, both share a predicted usefulness likelihood of 1. For each resolution, the raw classification data from Part 1 can be seen in inset four panels of Figure 3C. The solid lines show the logistic regression model and the dashed lines show the 95% confidence intervals around the fit.

### 3.2 Part 2: Is reconstruction error acceptable?

Next we investigate the effect of deep learning enhancement on image classification. The inter-rater reliability for the classification of enhanced images in Part 2 shows slight agreement between evaluators 1 and 2 (K = 0.15), fair agreement between evaluators 2 and 3 (K = 0.24), and moderate agreement between evaluators 1 and 3 (K = 0.48). Figure 4A shows a side by side comparison of ground truth (left column) with corresponding enhanced images (right column). Note the subtle errors in brain and CSF locations in the top right image and the more substantial errors in the lower right image. Regardless of these spatial errors, CNR is significantly increased by the enhancement network, as shown in the plot in Figure 4B where average CNR of test images at each network location are shown before and after enhancement using the logistic models developed in Part 1. These data predict very high usefulness likelihood for enhanced images based on increased CNR. The table in Figure 4C shows that while the Part 1 classification of enhanced images does closely follow the prediction of high usefulness likelihood, re-classification of enhanced images in Part 2 reveals that many enhanced images contain errors that are not clinically acceptable. We use an additional classification of Misleading for these images (i.e. images that were deemed Useful in Part 1, but had unacceptable errors in Part 2).

Since the logistic models developed in Part 1 do not describe the Part 2 classification, a new logistic regression model was constructed for Part 2 with pre-enhancement noise and contrast of images as predictors. Only contrast showed significance (Figures S13 and S14) so noise was removed from the model. In order to compare the usefulness likelihood of a degraded image (Part 1) with the risk of misleading errors in an enhanced image (Part 2), an additional logistic regression model with CNR as the predictor was computed based on Part 2 classification (Figure 4D). Risk of misleading results is calculated to be 1 minus the usefulness likelihood of the enhanced images based on a univariate logistic regression with CNR prior to enhancement as the predictor. As CNR increases, a 128×128 image is more likely to be useful in its degraded state (left vertical axis) and less likely to be misleading if enhanced (right vertical axis). Note that there exists no CNR value for which there is low usefulness likelihood of the degraded image *and* low risk of generating a misleading image through enhancement.

## 4 Discussion

### 4.1 Utility of Low CNR Images

The image quality threshold required for treatment planning of hydrocephalus is significantly lower than the quality typically provided by CT or high-field MRI imaging systems. The results in Figure 3B-C can be viewed in several different ways. CNR is a comparison between the signal-to-noise ratio of two regions of interest. This implies that the true limiting factor of image quality is per voxel signal-to-noise, for which high-field MRI has an inherent advantage over low-field MRI. However, Figure 3B-C suggests that there are options for using low CNR or low resolution images that may be advantageous. For a high-field system imaging infant hydrocephalus, a short scan time is desirable, in which case resolution and signal-to-noise can be traded for a faster scan. Alternatively, in the resource limited setting of an LMIC, a low-field MRI system has the potential to provide equivalent diagnostic information at a significant reduction in cost and complexity. The trade-off for this low cost and complexity is lower signal-to-noise and interpretability. It is the interpretability that sets the threshold for the lower bound of signal-to-noise.

The usefulness likelihood for the 3T (CNR = 13) and 0.05 T (CNR = 4) MRI images without deep learning enhancement featured in Figure 1 are indicated in Figure 3C. Although the visual quality of the two images is strikingly different, they are predicted to have the same utility for hydrocephalus treatment planning.

To put this in the context of global sustainability, the acquisition cost of the 0.05 T system used for producing the image in Figure 1B is less than $20,000 USD. A 3T system costs at least an additional $2.8 million USD (excluding siting, maintenance, and consumables) and it can provide over three times the CNR (Figure 1A). However, for the cost of a single 3T system, 150 low-field MRI systems could be placed throughout the region, providing increased access to the hydrocephalus patient population without compromise in diagnostic utility.

In addition to being a substantial global health need for children’s medicine, hydrocephalus is also an exceptionally straightforward technical challenge for low-field MRI systems. In the vast majority of hydrocephalic children, there is no need to differentiate contrast within the brain parenchyma for diagnosis, triage, monitoring, or treatment planning. For MRI the signal strength from the water-based CSF is the strongest signal within the head. Although our results support substantial utility from images with reduced quality in hydrocephalus management, more complex diagnostic and treatment decision-making in other diseases will pose additional challenges to such technologies.

### 4.2 Enhanced Images: Benefit or risk?

Image enhancement appears to perform exceptionally well based on Part 1 data, as shown in Figure 4A where even the worst network locations are more than 85% likely to be rendered useful. However, data from Part 2 reveals that enhancement yields images that appear useful, but in fact would mislead treatment decisions due to unacceptable errors in brain and CSF location. Subtle features in the configuration of the CSF spaces, such as increased rounding of brain ventricles, are important signs of increased intracranial pressure suggesting that surgery might be required to improve CSF diversion through a shunt or endoscopic fenestration. If features such as these are a product of the enhancement network and not indicative of the true condition of the disease, clinicians may be led to make poor treatment decisions.

The key difference in using a degraded image versus its enhanced counterpart in a clinical setting is the source of risk. A degraded image is either useful or it is not - the risk of using it to diagnose or treat disease rests with the judgement of the clinician. Enhanced images in this study yield useful looking images 99% of the time, however 75% of these images are shown to have uncertain utility or to be misleading after comparison with ground truth. The risk of enhancement arises from the black box of the deep learning network. Furthermore, as shown in Figure 4D, there is never a CNR for which there is low risk of producing a misleading image and low usefulness likelihood of the degraded image without enhancement. For example, a 128×128 degraded image with with modest CNR yielding 75% usefulness likelihood still has a 14% chance of producing a misleading image through enhancement. Enhancing highly degraded images can improve the usefulness likelihood, but with substantially increased risk of misleading results. We find no scenario in which enhancement is safely beneficial. Note also that the CNR of the 0.05 T system studied had a very high useful likelihood and would not have required enhancement. Yet acceptance of such unenhanced images as shown in Figure 1B would constitute a cultural shift in current standards of diagnostic acceptability.

Machine learning can generate attractive images from patterns with highly degraded information content. Philosophically, a learning library of other patient images enables utilization of information not present in the individual case undergoing enhancement. Such learned information brought to a new case image can be clinically misleading. This is a very different situation from machine learning faces or objects, or diagnosis classification from images, where there is only one correct match and the information required is already in the learning library. Hydrocephalus, as in so many other pathological conditions, tends to produce a unique structural pattern for each patient. For machine learning, automating the choice of a diagnosis is therefore very different from reconstructing an unknown unique architecture. This fundamental issue implies that while this study only employed one learning network architecture, this risk likely exists in other machine learning strategies and great care should be taken when employing these methods for anatomic reconstruction. A challenge for the machine learning community working with low-resolution and low-contrast images is to improve interpretation while minimizing risk of clinical errors.

### 4.3 Limitations

This study has limitations. Only three experts participated in the assessment. The single central slice from the image stack was chosen to demonstrate image quality and enhancement. Only image quality concerns inherent to low-field systems such as noise and contrast were considered, while distortions in the low-field image were not. Only one deep learning network architecture was employed and the number of training samples was relatively low (80). However, the size of available image archives is typically not large for diseases unique to LMIC such as post-infectious hydrocephalus in sub-Saharan Africa. A more complex machine learning strategy could incorporate a 3D array of connected slices for enhancement and clinical review.

Although motivation for this study stems from the advent of clinical low-field MRI as a tool for hydrocephalus treatment planning, the work was conducted with CT images. In high-resource settings where low-field MRI is being deployed (such as intensive care units), CT remains the high-resolution alternative of choice [11]. CT is the high-resolution modality most available in LMIC, and currently the only available repository of postinfectious hydrocephalus images where low-field MRI will soon be deployed. Note that we argue the potential benefits of low-field MRI using only one example image in Figure 1B. This can be extended in the future as reliable low-field MR image repositories become available. We anticipate that this quantifiable measure of CNR between brain and CSF will be generalizable to MRI at various field strengths as well as other CT studies of infant hydrocephalus treatment planning. Further evaluation will be necessary to determine whether CNR proves an important classifier for other conditions that may have more stringent image quality requirements.

## 5 Conclusion

The true value of a clinical medical image is in the treatment guiding information that it conveys to those providing care and in the patient outcomes that result, rather than its visual appeal. We have shown that lower quality images that are not customarily considered acceptable can be useful in planning hydrocephalus treatment. In addition, image resolution and contrast-to-noise ratio of brain and CSF predict the likelihood of a useful image for hydrocephalus treatment planning. Although deep learning can dramatically improve the visual quality of a highly degraded image, there is a substantial risk of misleading results, and algorithmic guidelines should be developed to avoid structural alterations which are potentially hazardous to clinical interpretation. At present, the most valuable low-resolution images may be less enhanced versions that maintain the structural details undistorted by excessive deep learning processing; indeed, emerging low-field MRI technologies are capable of producing useful images for hydrocephalus treatment planning without enhancement. Our findings advocate for new standards in assessing the cost-effectiveness of sustainable imaging technologies that can broaden global access to diagnostic imaging, and a reconsideration of acceptable image quality for clinical use.

## Data Availability

All data from the trial will be made available to qualified researchers upon request. The original patient images are not available to share as these form part of an ongoing clinical trial followup.

## 6 Acknowledgements

Supported by US National Institutes of Health grant R01HD085853. ClinicalTrials.gov registration number NCT01936272.

## 7 Supplementary Methods

### 7.1 Part 1 Instructions and Examples

During Part 1 of the assessment, you will be shown CT images of infant hydrocephalus. The images shown will be from a variety of different quality levels.

Your task will be to assess whether you think the images shown could be USEFUL to start treatment in a low resource setting.

To make a selection of an image that you think IS useful click the button below the corresponding image.

After you have reviewed all four images and made the desired selections, click the SUBMIT button to log the data and generate four new images.

Thank - you for your participation.

**Figure S1:**
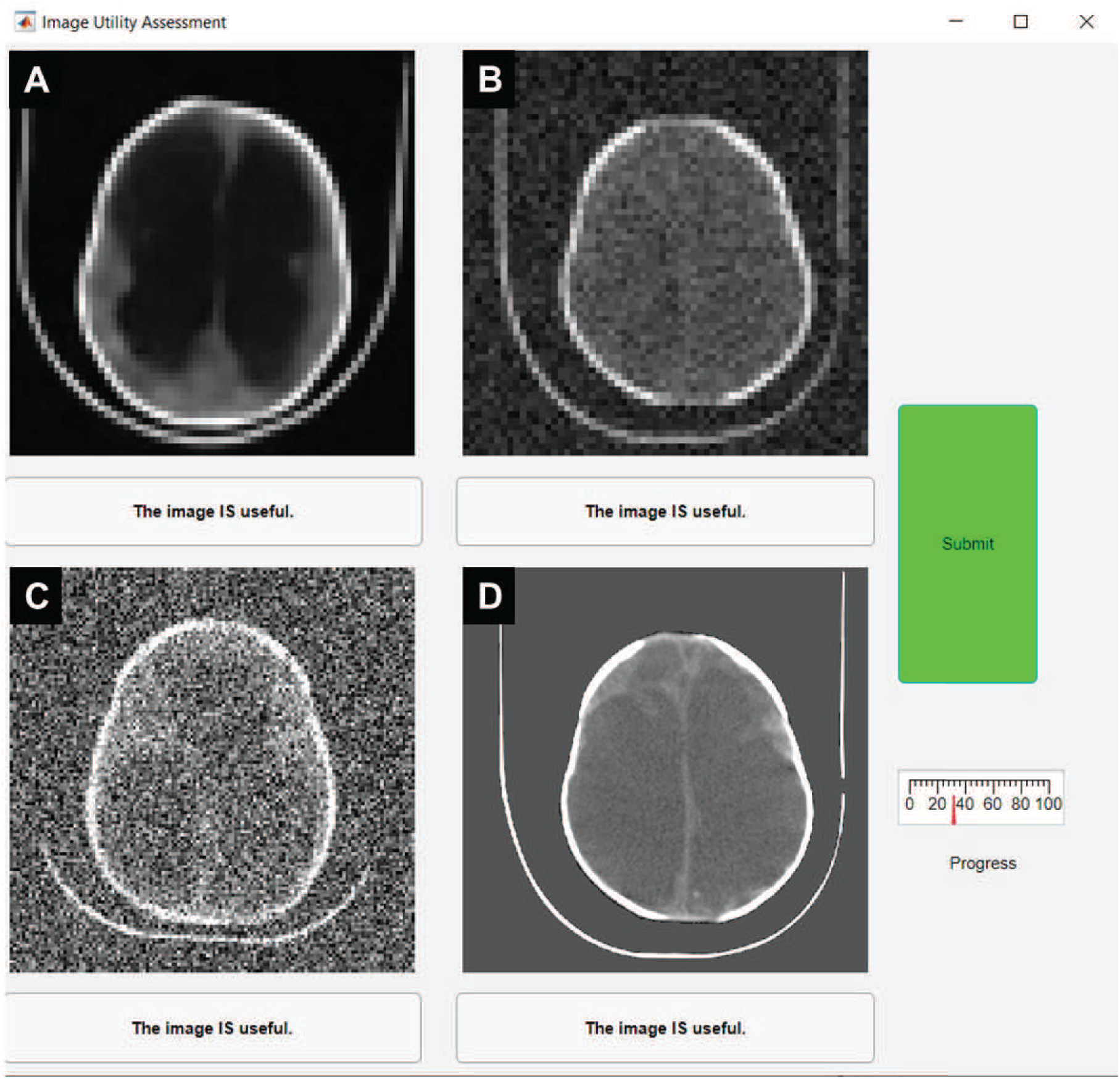
An example panel from Part 1 of the assessment showing A) An enhanced image with resolution: 64×64, characteristics before enhancement of noise variance: 0.006 and contrast loss: 60%, and classification: Useful; B) Image with resolution: 64×64, noise variance: 0.001, contrast loss: 71%, and classification: Not Useful; C) Image with resolution: 128×128, noise variance: 0.03, contrast loss: 0%, and classification: Uncertain; D) Image with resolution: 512×512, noise variance: 0, contrast loss: 42%, classification: Useful.

**Figure S2:**
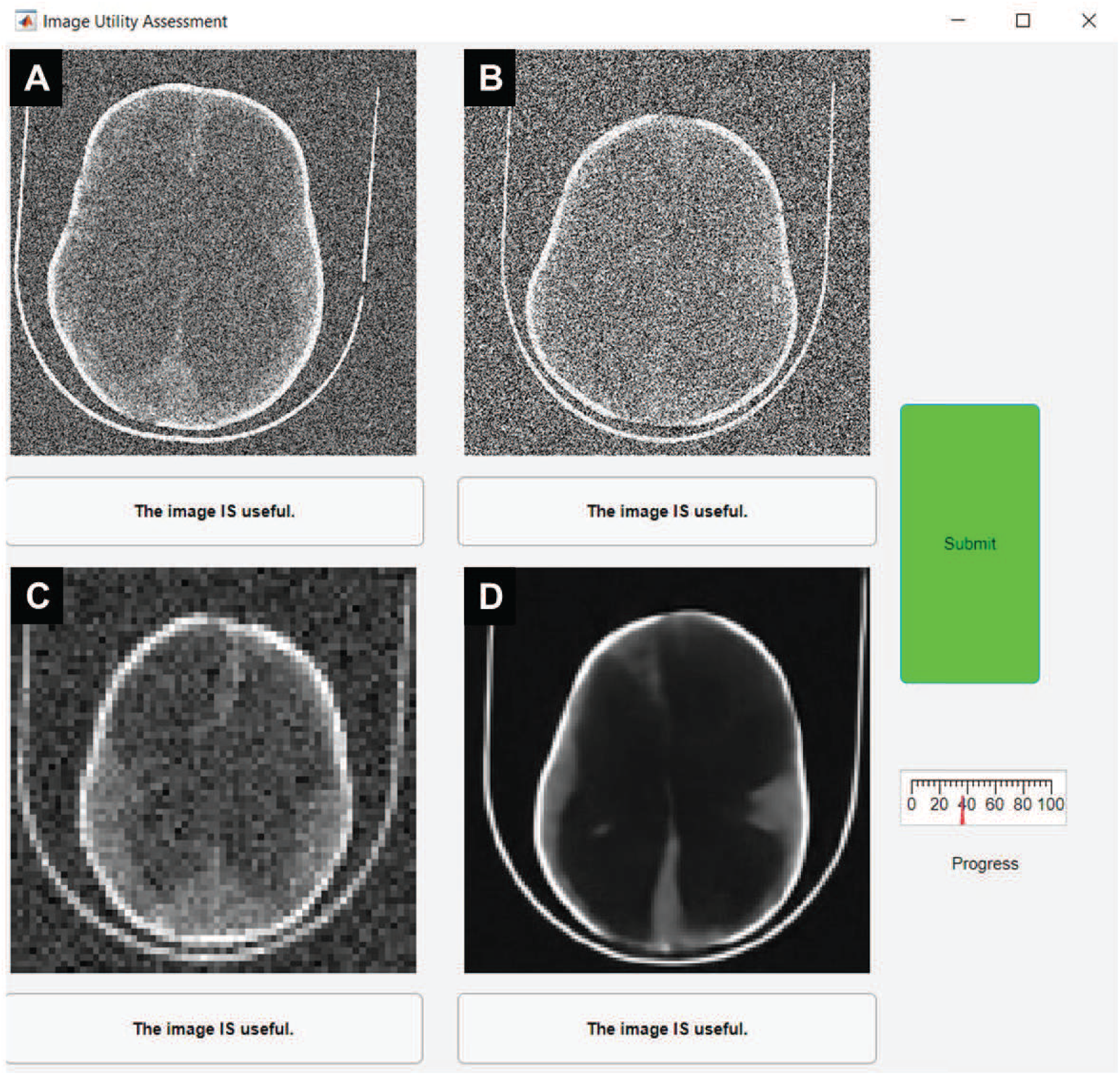
An example panel from Part 1 of the assessment showing A) Image with resolution: 512×512, noise variance: 0.05, contrast loss: 35%, classification: Uncertain. B) Image with resolution: 512×512, noise variance: 0.1, contrast loss: 64%, classification: Not Useful. C) Image with resolution: 64×64, noise variance: 0.003, contrast loss: 14%, and classification: Useful; D) An enhanced image with resolution: 128×128, characteristics before enhancement of noise variance: 0.03 and contrast loss: 60%, and classification: Useful.

### 7.2 Part 2 Instructions

A selection of images shown during Part 1 of the assessment were low quality images enhanced using library learning. You chose some of these images for the “useful” category. All enhanced images contain errors.

In Part 2 of the assessment, you will be shown pairs of images, two at a time. Images on the left will be high-resolution “ground-truth” slices of a particular case and images on the right will be the enhanced version of the degraded image for the same slice and same case.

Would the error affect treatment in terms of risks and benefits? You will be asked to indicate whether the error is acceptable by pressing the button associated with each pairing.

**Figure S3:**
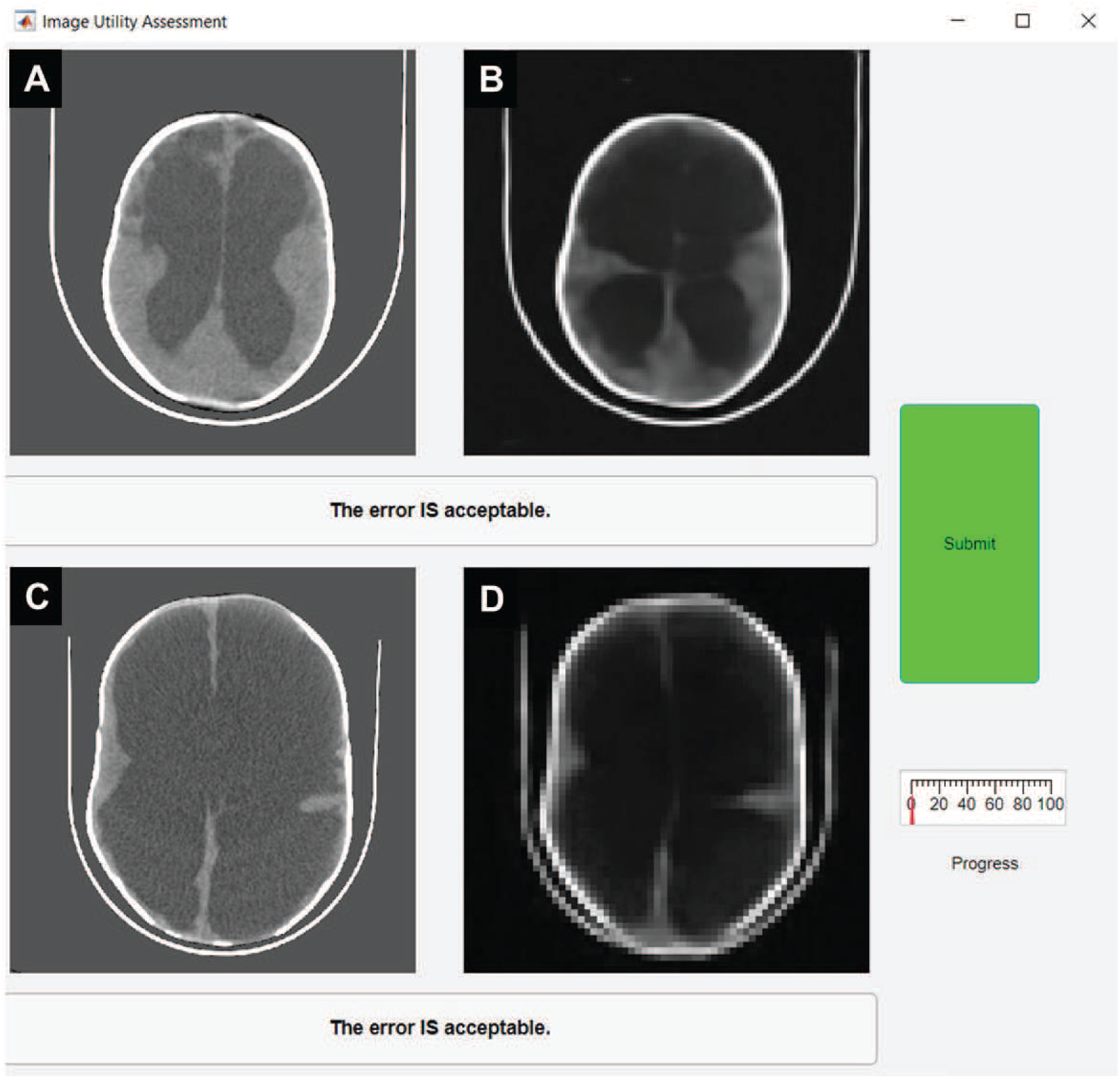
An example panel from Part 2 of the assessment showing A) A ground-truth 512×512 CT image, and B) the enhancement of a degraded version of (A) with resolution: 128×128, characteristics before enhancement of noise variance: 0.05 and contrast loss: 60%. Part 1 classification: Useful. Part 2 classification: Not Useful. C) A ground-truth 512×512 CT image, and D) the enhancement of a degraded version of (C) with resolution: 64×64, characteristics before enhancement of noise variance: 0.003 and contrast loss: 60%. Part 1 classification: Useful. Part 2 classification: Useful.

**Figure S4:**
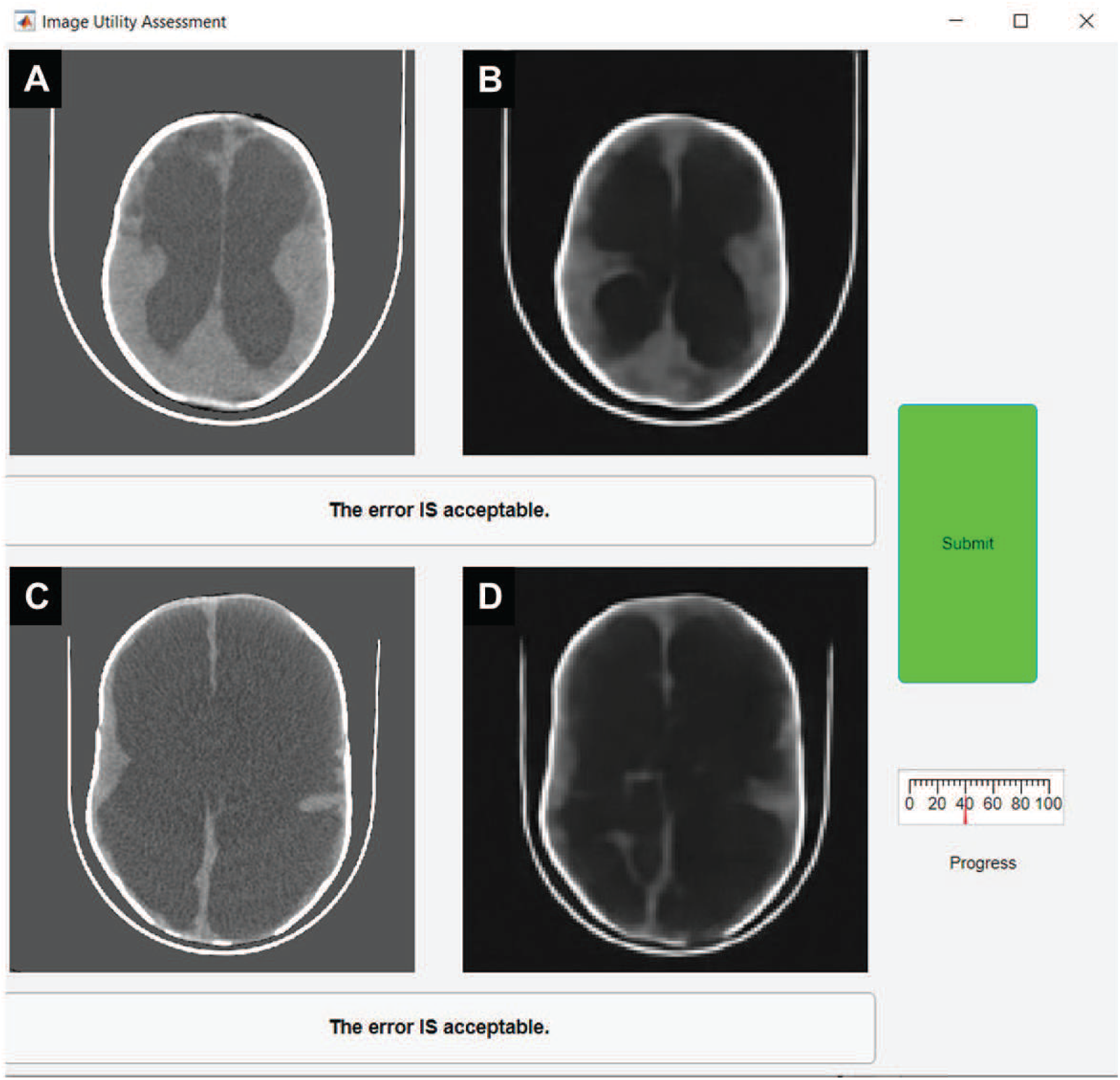
An example panel from Part 2 of the assessment showing A) A ground-truth 512×512 CT image, and B) The enhancement of a degraded version of (A) with resolution: 128×128, characteristics before enhancement of noise variance: 0.02 and contrast loss: 60%. Part 1 classification: Useful. Part 2 classification: Uncertain. C) A ground-truth 512×512 CT image, and D) the enhancement of a degraded version of (C) with resolution: 128×128, characteristics before enhancement of noise variance: 0.02 and contrast loss: 60%. Part 1 classification: Useful. Part 2 classification: Useful.

### 7.3 CT Images

There were 90 CT images used in this study. Of these, 10 were randomly selected as test images and are shown in Figure S5. All degradations and enhancements presented in the image utility assessment were performed on these 10 images. The remaining 80 images were used as a learning library for the deep learning enhancement networks.

**Figure S5:**
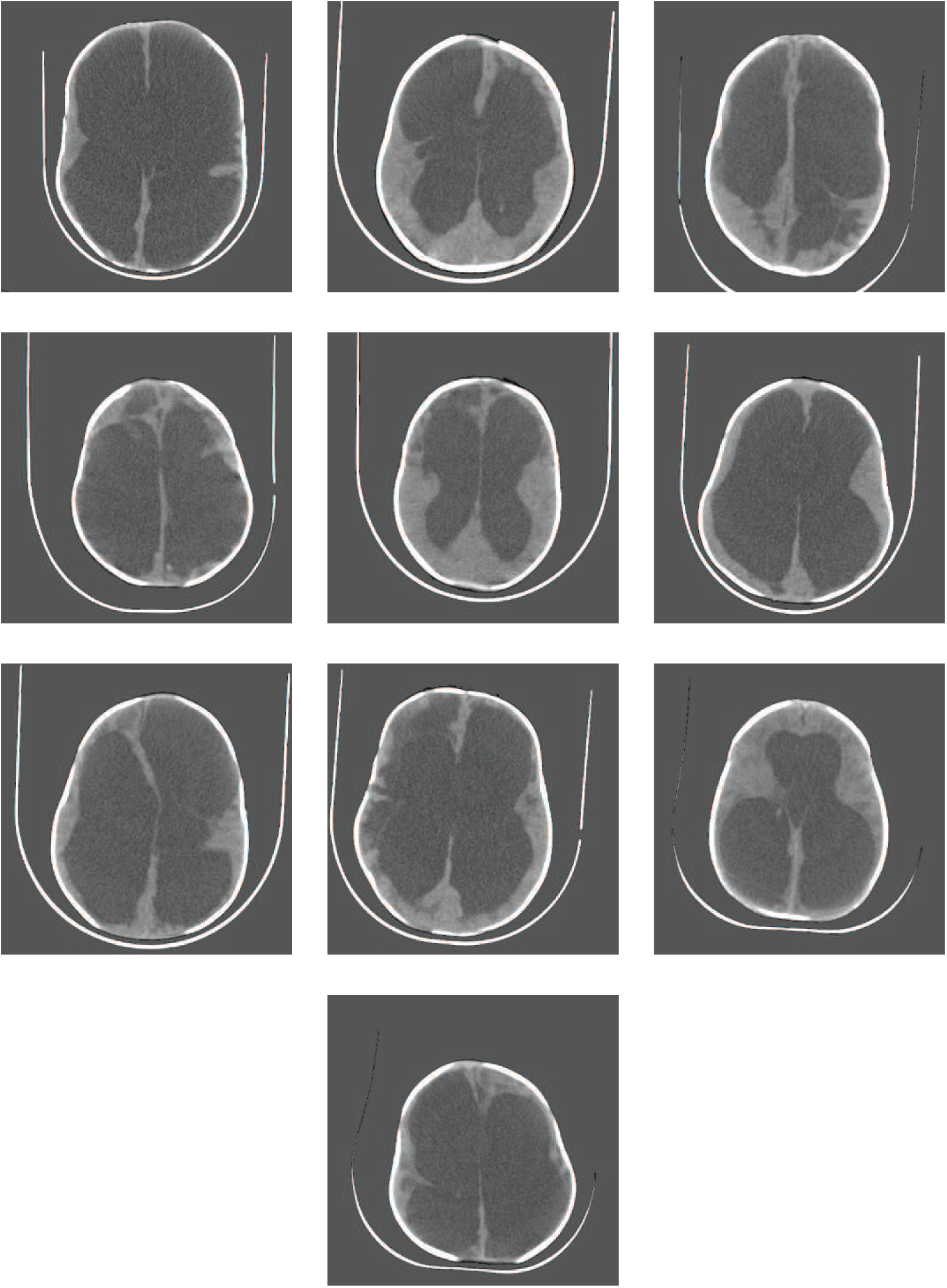
The 10 randomly selected central slice test images chosen for the image utility assessment.

### 7.4 Histogram Compression

Prior segmentation of brain and CSF for the 10 test slices is necessary in order to perform this algorithm. Figure S6 shows an example of the histogram compression technique on a test image. In this figure, the original image is shown at the top next to the histogram of gray-scale values for CSF and brain. This is compared to histogram overlapping (middle) in which the entire histogram for brain is translated so that it overlaps the histogram for CSF. While this does significantly reduce contrast between brain and CSF, the displacement of histogram values for pixels bordering brain and CSF creates a bright line at the interface of the tissues (gray scale values of CSF pixels bordering brain are translated to higher gray scale values and gray scale values of brain pixels bordering CSF are translated to lower gray scale values). This is not desirable as it highlights the boundary. Histogram compression (bottom) reduces contrast without highlighting boundaries, which simulates an image generated with little contrast between tissues.

**Figure S6:**
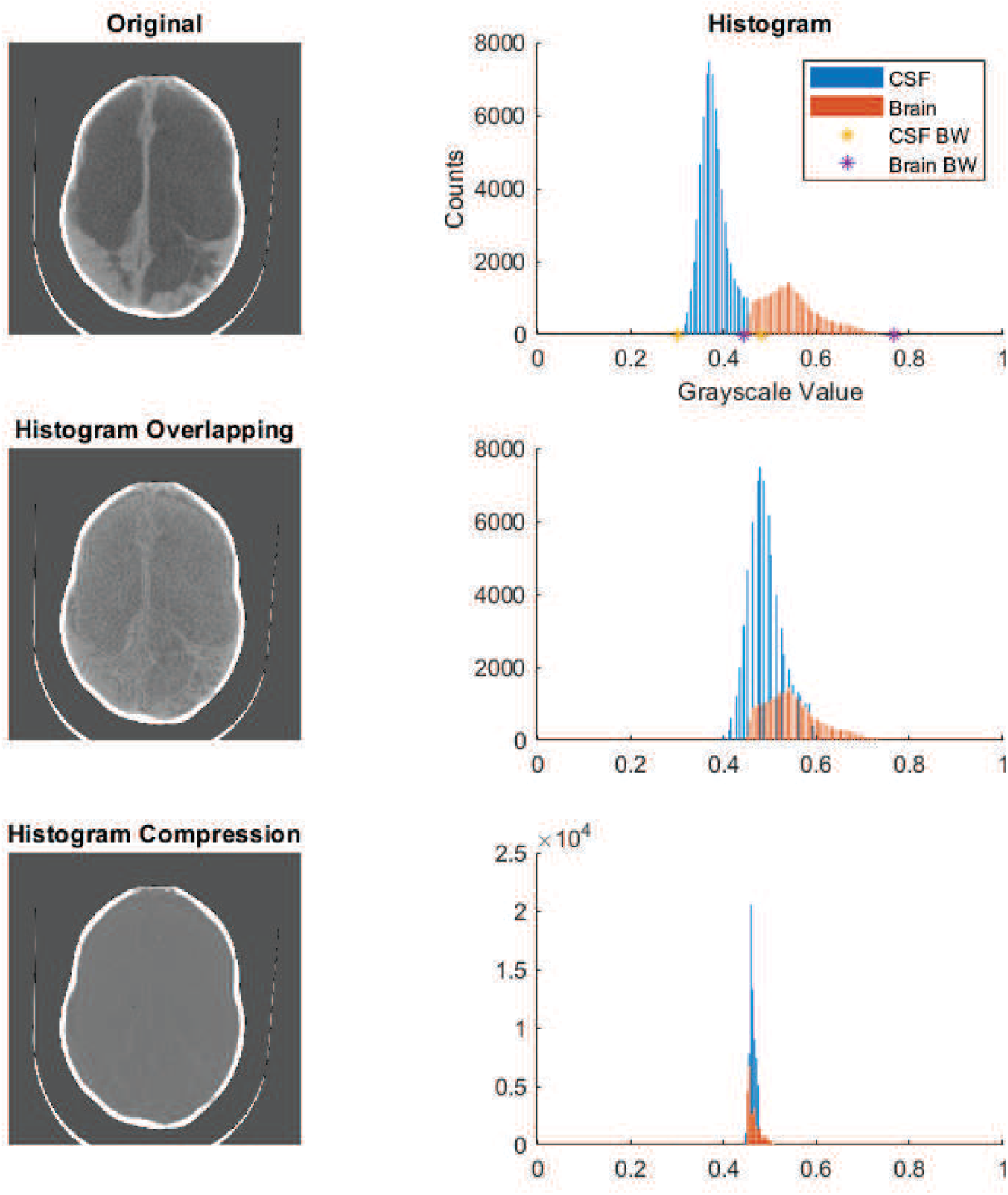
Example of the histogram compression algorithm. The original image (top) is shown next to the histogram of brain and CSF gray-scale values. The yellow ‘*’ shows the extents of CSF values and the purple ‘*’ shows the extent of brain values. Histogram overlap (middle) creates bright lines at the boundary between brain and CSF. Histogram compression (bottom) reduces contrast and eliminates the bright lines between brain and CSF by preserving the natural gray-scale boundary on the histogram.

The histogram compression algorithm works by first finding the maximum and minimum gray-scale values in both brain and CSF, as shown by the yellow and purple asterisks in Figure S6 at the top. From these points, the median gray-scale value is calculated for brain and CSF and the distance between the medians is measured. Then, histogram values from the smallest CSF bins are moved to the next largest bin and histogram values from the largest brain bin are moved to the next smallest bin. The mean is measured again and the fractional change in the distance between medians is considered the contrast adjustment. The process is iterated to the desired contrast level. A contrast reduction of 0 corresponds to the distance between bins that exists in the native image without histogram compression. A contrast reduction of 1 corresponds to the case where medians of brain and CSF bins completely overlap (i.e. contrast has been reduced to the point where the difference between tissues can no longer be separated). This process was inspired by the well-established algorithm of histogram equalization for increasing contrast in an image, in which the histogram of an image is spread across as many gray-scale values as possible [41, 42].

### 7.5 Noise Degradation

The maximum noise added in parameter space was resolution specific. This is because noise variance has a higher impact on lower resolution images since, in a sense, more information is lost per corrupted pixel. The resolution specific levels were chosen experimentally by observing the quality of the output images and attempting to choose a level that would demonstrate the threshold between useful and not useful images. In parameter space, each resolution was normalized to its maximum noise value and 20 levels between 0 (no noise added) and 1 (maximum noise added) were used.

### 7.6 CNR

Since contrast, noise, and resolution are the factors in the image parameter space, contrast-to-noise ratio (CNR) was selected as the quantitative measure of degraded image quality. Average CNR across parameter space was generated by measuring the CNR between brain and CSF for each of the 90 images (test + library) at each point in parameter space (1600). Then the mean CNR was taken across the images at each point in parameter space. This allows for mean CNR to be mapped into parameter space for further analysis. Additionally the CNR of each individual test image at the 420 test locations was calculated. CNR for each image was calculated using the following:

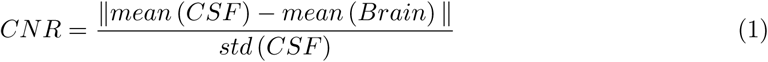

where *CSF* and *Brain* represent the gray-scale values found in the segmented regions of CSF and Brain. Typical CNR calculations might use the standard deviation of signal in a region of the image without tissue to estimate noise standard deviation of the entire image. Given the variable dimensions of the brains in the image set and the existence of the head rest in most images at variable locations, it was difficult to choose a good location outside of the brain that would represent purely noise space. Since noise in CT is consistent across the entire image and since CSF should show a relatively homogeneous signal, the standard deviation of CSF values in the image was chosen to model for noise standard deviation for the entire image.

### 7.7 Enhancement Networks

Deep learning networks were trained at locations in parameter space that were likely to be regions that are not useful, as can be seen in the left panel of Figure S7. The axes of Figure S7 are normalized, however the table in the right panel Figure S7 shows the true noise variance added and contrast reduction used for 64×64 and 128×128 resolutions.

**Figure S7:**
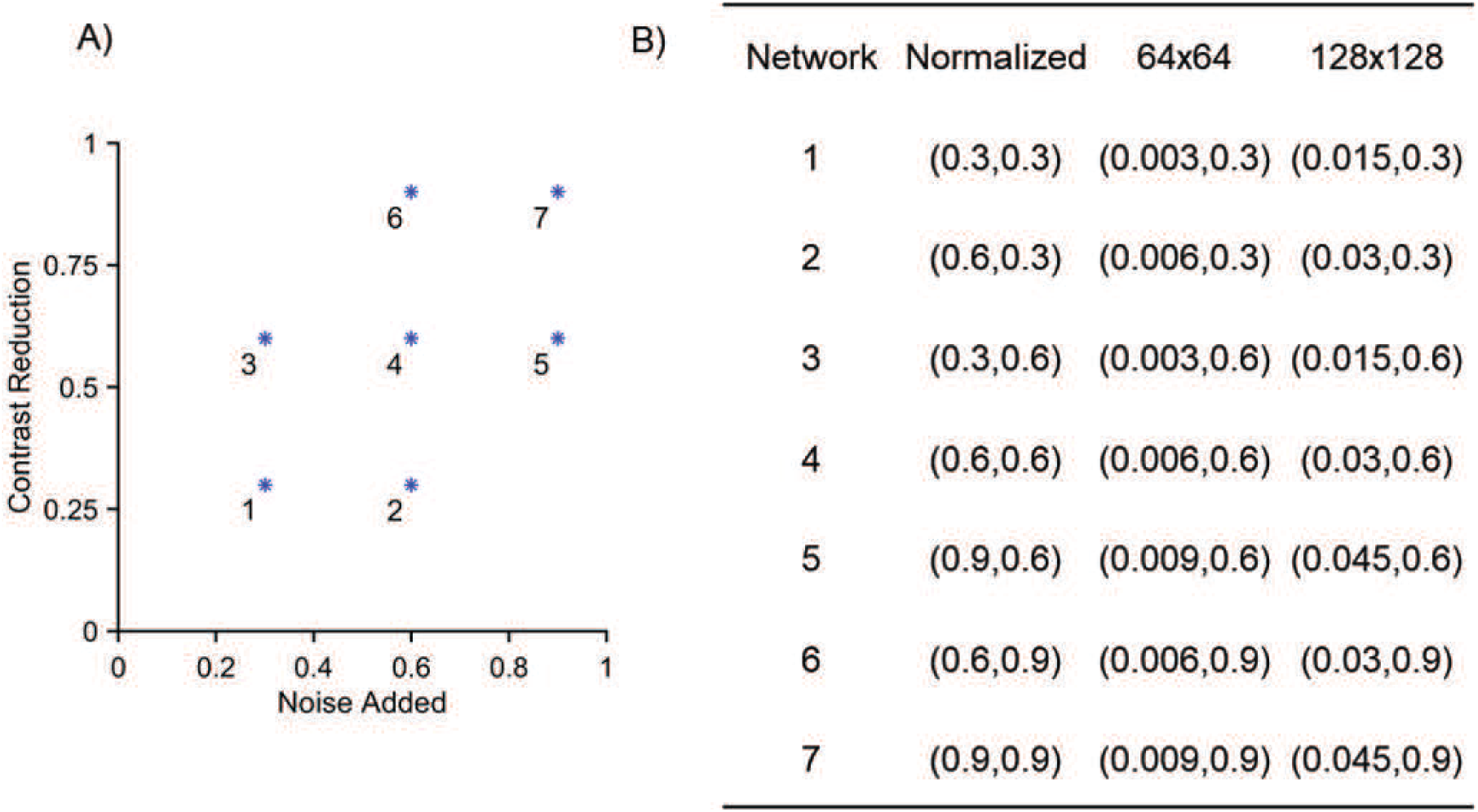
Locations of enhanced networks on the noise added vs. contrast reduction parameter plane. The locations are numbered 1 to 7 with 1 being the least degraded. Actual noise and contrast reduction values for each network are shown on the right by resolution.

### 7.8 Inter-Rater Reliability

Cohen’s Kappa (K) provides a method for comparing the agreement between two evaluators with the agreement expected by chance. It is appropriate for nominal data and fully crossed study designs like the present study. A common guideline for interpreting Cohen’s Kappa is that Kappa values between 0 and 0.2 suggest slight agreement, 0.21 to 0.40 suggests fair agreement, 0.41 to 0.60 suggests moderate agreement, 0.61 to 0.80 suggests substantial agreement, and 0.81 to 1 suggests nearly perfect agreement [40]. The extension of Cohen’s Kappa described in [39] accounts for the existence of prevalence through a Prevalence Index (PI) and bias in the data through a Bias Index (BI). Prevalence refers to the disproportionate occurrence of “yes” responses, which is known to reduce the Kappa measure. The prevalence index ranges from -1 to 1 with negative values indicating prevalence for negative responses. Bias refers to disproportionate “yes” responses between evaluators which is known to increase the Kappa measure. The Bias index ranges from 0, indicating no bias, to 1 indicating complete bias.

We divided this analysis into three parts: 1) The classification of degraded images from Part 1, excluding enhanced image classification, 2) the classification of enhanced images from Part 1, and 3) the re-classification of enhanced images compared to ground truth in Part 2 of the assessment. We began by constructing tables of agreement between each evaluator (i.e. agreement between evaluator 1 and 2, between evaluator 1 and 3, and between evaluator 2 and 3 for each of the three divisions) and the associated PI, BI, and K values.

## 8 Supplementary Results

### 8.1 Part1

The inter-rater reliability for degraded image classification and Part 1 enhanced image classification can be seen in Figure S8 and Figure S9, respectively. Figure S8A shows that there is fair agreement between evaluators 1 and 2 (K_12_=0.33) with bias (BI_12_=0.37) and a prevalence for negative responses (PI_12_=-0.22). Figure S8B shows that there is almost perfect agreement between evaluators 2 and 3 (K_12_=0.94) with almost no bias (BI_12_=0.02) and a prevalence for negative responses (PI_12_=-0.57). Figure S8C shows that there is fair agreement between evaluators 1 and 3 (K_12_=0.34) with bias (BI_12_=0.34) and a prevalence for negative responses (PI_12_=-0.20).

**Figure S8:**
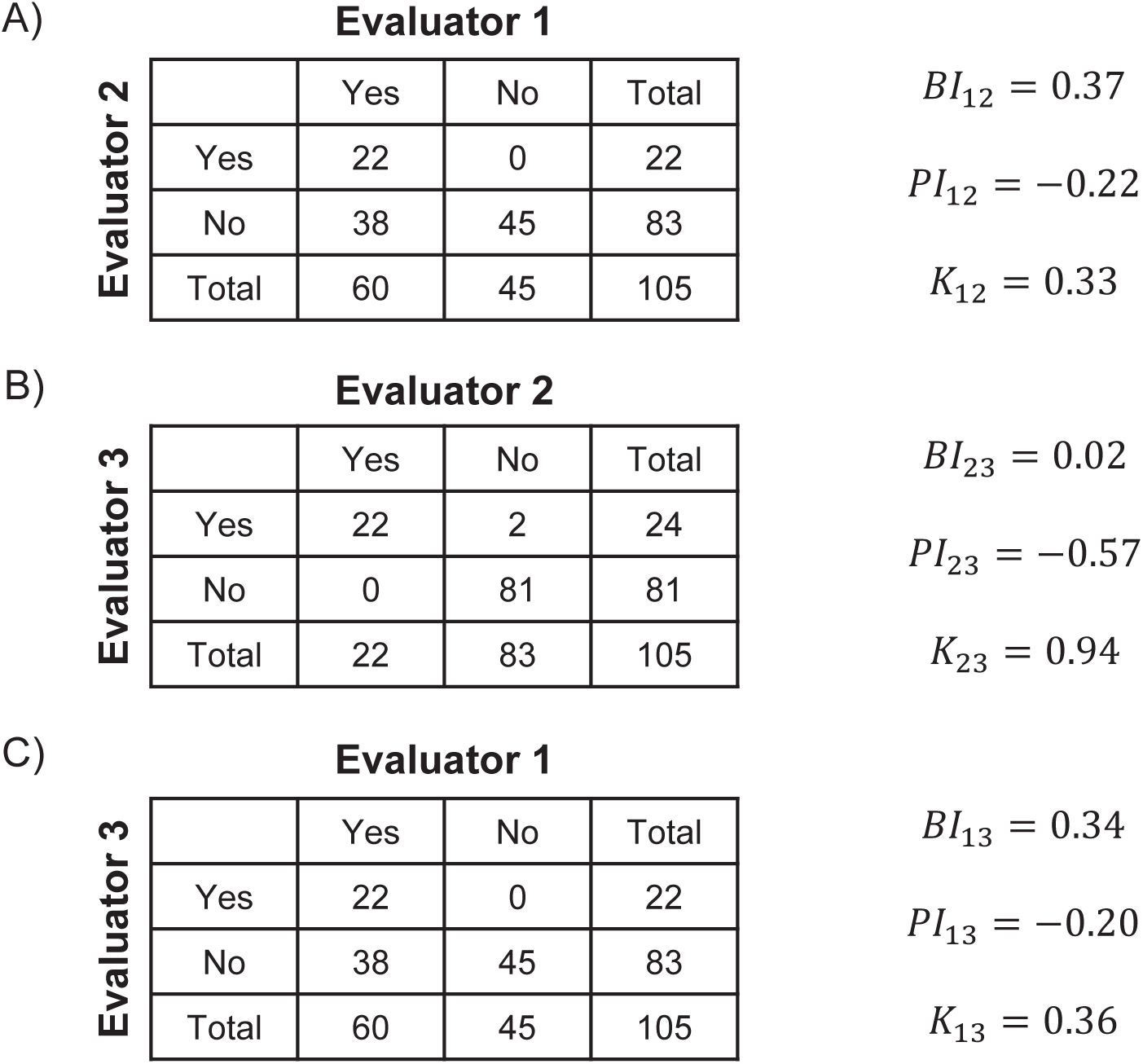
Agreement between each evaluator for classification of Part 1 degraded images is shown along with the Bias Index (BI), Prevalence Index (PI), and Kappa value (K). A) shows the agreement between evaluators 1 and 2. B) shows the agreement between evaluators 2 and 3 and C) shows the agreement between evaluators 1 and 3.

The table in Figure S9A shows that there was total agreement between evaluators 1 and 2 and that there was perfect prevalence of positive responses (PI_12_=1). In cases such as this, it is not possible to calculate a K value (K_12_=N/A). Figures S9B and C shows that there was nearly perfect agreement between evaluators 2 and 3 and evaluators 1 and 3. This is because evaluator 3 was the only evaluator to classify any enhanced images as “not useful” in part 1. In both cases, the prevalence index is nearly 1 which indicates the calculated value of K cannot be interpreted correctly [39] and we must consider the agreement table rather than Cohen’s Kappa to determine that there is a high level of agreement.

**Figure S9:**
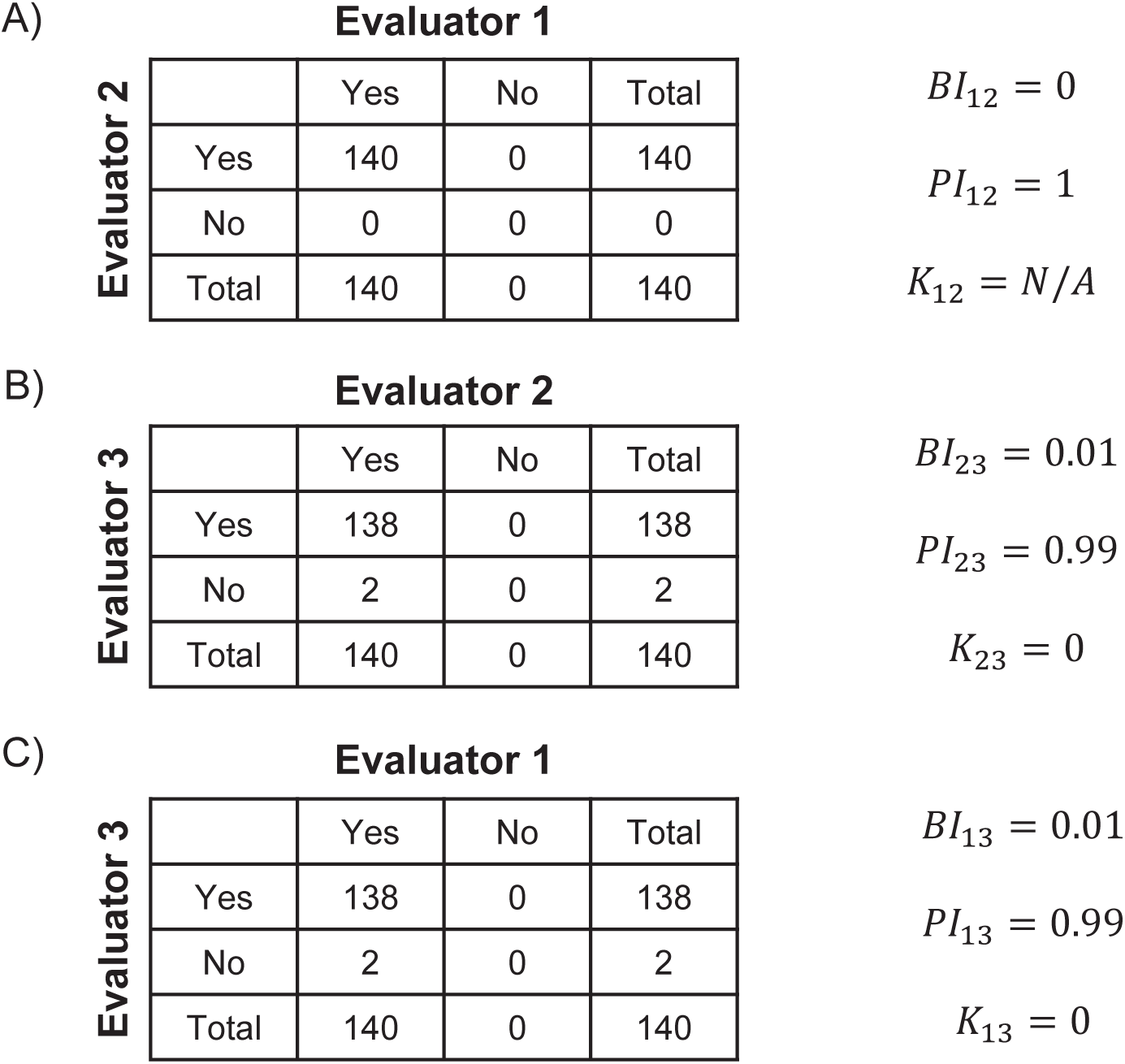
Agreement between each evaluator for classification of Part 1 enhanced images is shown along with the Bias Index (BI), Prevalence Index (PI), and Kappa value (K). A) shows the agreement between evaluators 1 and 2. B) shows the agreement between evaluators 2 and 3 and C) shows the agreement between evaluators 1 and 3.

The degraded image classification data is plotted in Figure S10 separated by resolution. The contrast reduction scale is common across all resolutions and the noise variance added is resolution specific, as described in methods. Lines of constant mean CNR are plotted for each resolution shown. Triangles in Figure S10 show datapoints in which all three experts agreed that the image was useful. Circles show datapoints in which 2 or only 1 expert(s) classified the image useful, and “X”s show datapoints in which none of the 3 experts classified the image useful. A logistic regression model was implemented using MATLAB, with contrast reduction and noise added as predictors, and classification as the response. The number of trials was set to 3 to account for responses from the 3 participants. The p-values following a deviance test for each resolution is as follows: p_32×32_ = 7e-6, p_64×64_ = 4e-27, p_128×128_ = 2e-17, p_512×512_ = 8e-32. The p-values indicate that the logistic regression models fit the data better than the null model. The null model is the average probability of a classification at a given resolution being useful. Lines of constant usefulness likelihood can be seen for each resolution. Values of average CNR across all images (10 test images + 80 learning library images) at each degradation point were also calculated and plotted as contours with a solid line.

**Figure S10:**
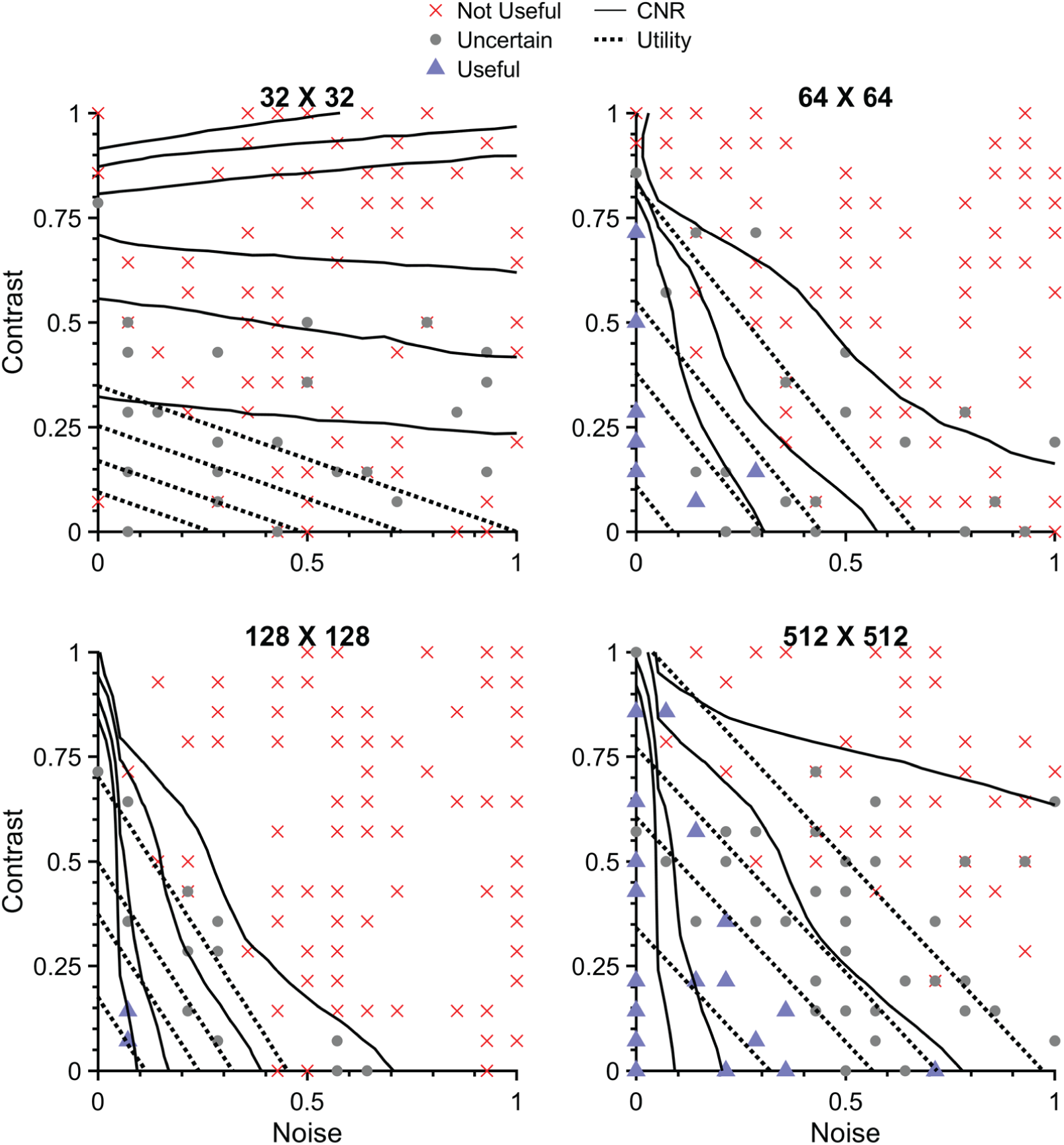
The combined classification of degraded images form the image utility assessment divided into 4 parameter planes of resolution 32×32, 64×64, 128×128, and 512×512. Lines of constant mean CNR are shown with the solid contour lines. Lines of constant usefulness likelihood as determined by the logistic regression model are also shown using the dotted lines. Higher usefulness likelihood is always closer to the origin.

As expected, 32×32 resolution offers very low likelihood of being useful. This is apparent in the raw data as well since there were no images that all three experts agreed to be useful. The other three resolutions do however show that a fair amount of degradation at low resolution has a high likelihood of being useful for planning hydrocephalus treatment. It is important to note that since the noise added axes are normalized to a different maximum value for each resolution the effect of resolution on usefullness likelihood is not visually evident in these plots. For example, the entire 0 to 1 range of normalized noise added for the 64×64 resolution is equivalent to the range 0 to 0.2 of noise added for the 128×128 resolution, or 0 to 0.07 in the 512×512 resolution. As resolution decreases, noisy images are less likely to be useful.

CNR appears to be an effective approximation of the likelihood lines in some cases. It makes intuitive sense that images which have been degraded by adjusting contrast and adding noise may be classified well by CNR. Figure S11 shows the receiver operating characteristic (ROC) curves for CNR and logistic regression as a classifier of useful images by resolution. ROC curves are generated by tabulating the true positive fraction (TPF) and false positive fraction (FPF) for a particular metric at all possible levels of classification. For example, when using CNR as a classifier, the maximum CNR value could be considered as the cutoff for classifying a useful image vs. a not useful image. All images with a CNR value at or above the maximum CNR would be classified as useful, and all images with a CNR below the maximum value would be considered not useful. This classification is compared with the actual, qualitative classification of the images from the image utility assessment and the TPF and FPF are calculated. The process can be repeated for each level of CNR down to the minimum generating a range of TPF and FPF pairs. These pairs create the ROC curve. The area under the curve (AOC) is good measure for comparison between the two methods and shows relative agreement within resolution. For both methods, classification is successful at higher spatial resolutions, with 32×32 being the poorest classification, as expected.

**Figure S11:**
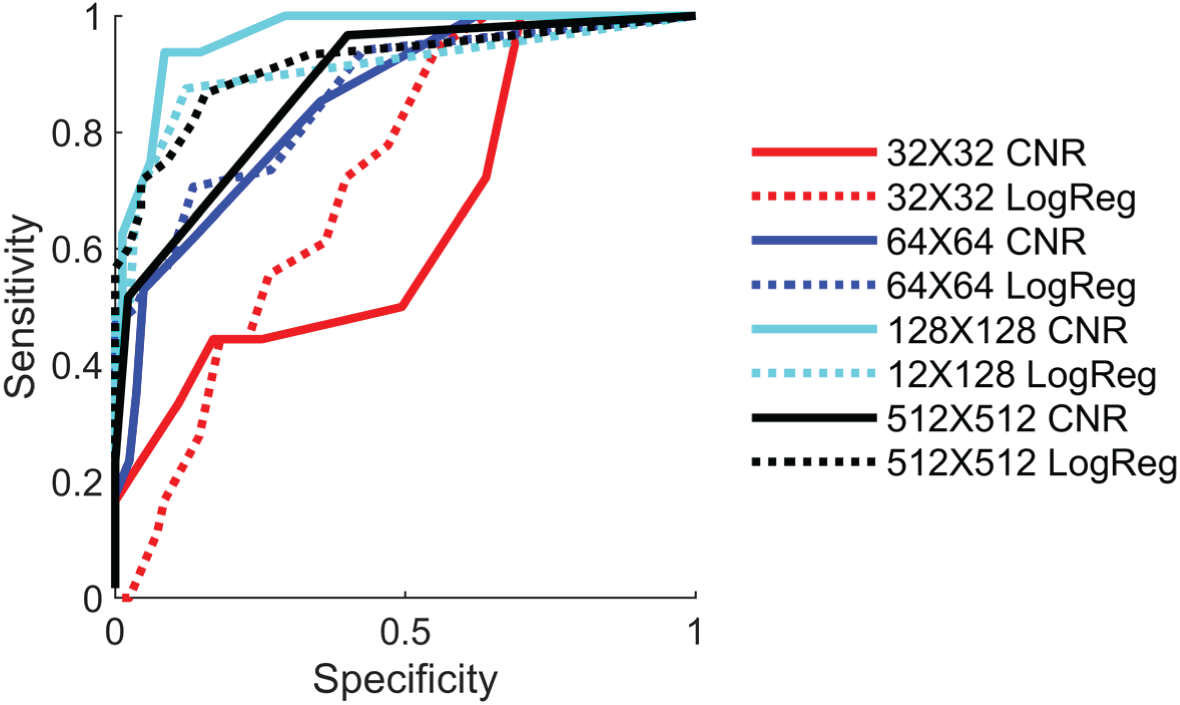
The ROC curves for CNR as a classifier of image utility compared to the logistc regression model as a classifier of image utility. For all resolutions the area under the curve is greater than 0.5.

It should be noted that while the two classification methods are comparable, CNR is an innate feature of the image and is more closely linked to the visual information that is important for hydrocephalus. Agreement of classification using CNR with logistic regression serves to strengthen the notion that CNR itself is an estimate of image utility.

### 8.2 Part 2

The inter-rater reliability for Part 2 classification of enhanced images can be seen in Figure S12. Figure S12A shows that there was slight agreement between evaluators 1 and 2 (K_12_=0.15) with bias (BI_12_=0.32) and a prevalence of negative responses (PI_12_=-0.29). Figure S12B shows that there was fair agreement between evaluators 2 and 3 (K_23_=0.24) with bias (BI_23_=0.39) and a prevalence for negative responses (PI_23_=-0.22). Figure S12C shows that there was moderate agreement between evaluators 1 and 3 (K_13_=0.48) with very little bias (BI_13_=0.07) and only slight prevalence for negative responses (PI_13_=-0.10).

**Figure S12:**
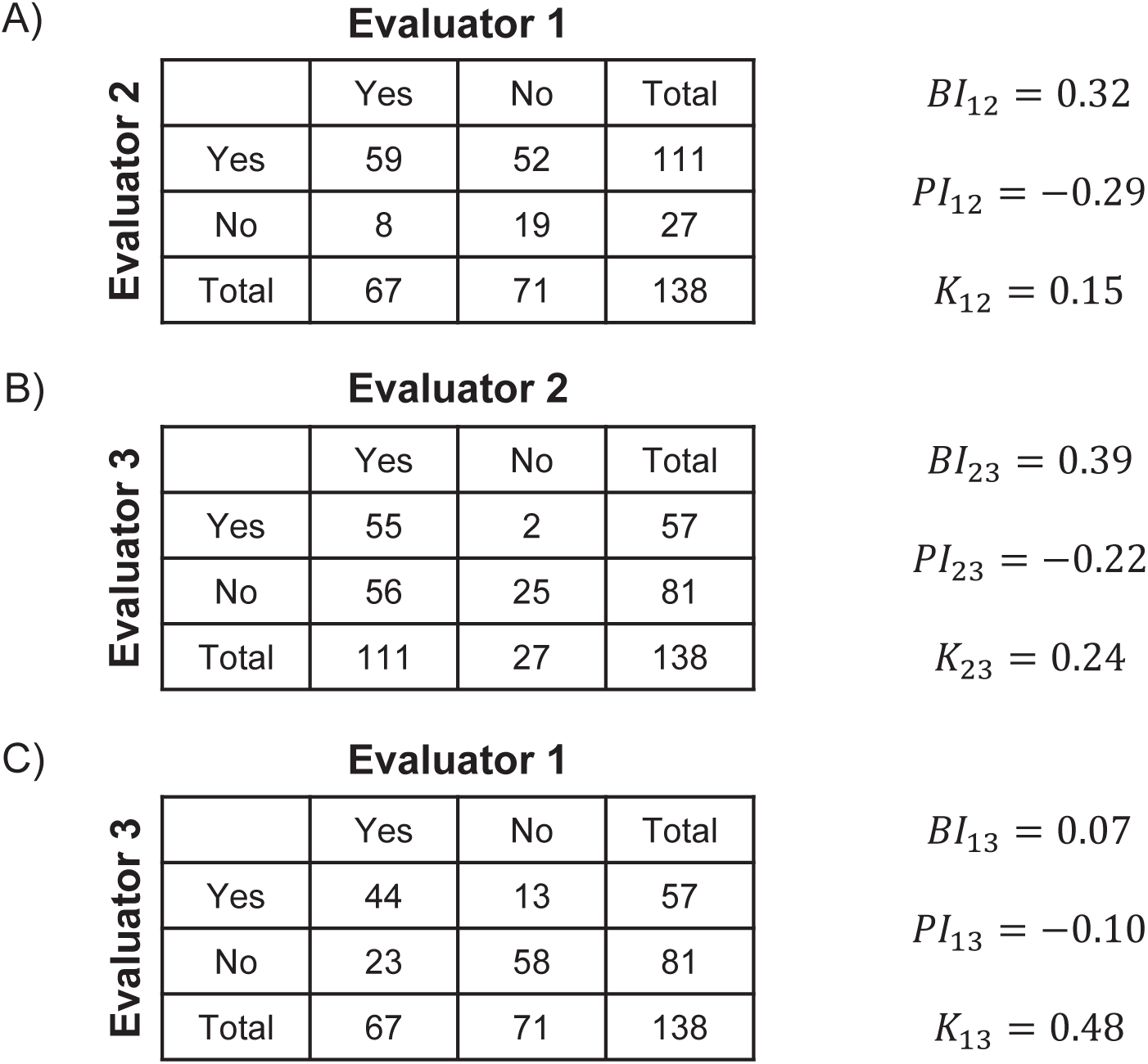
Agreement between each evaluator for classification of Part 2 enhanced images is shown along with the Bias Index (BI), Prevalence Index (PI), and Kappa value (K). A) shows the agreement between evaluators 1 and 2. B) shows the agreement between evaluators 2 and 3 and C) shows the agreement between evaluators 1 and 3.

The 10 test images were enhanced using the previously described Deep Learning network. Networks were trained at 7 locations for the resolutions 64×64 and 128×128. Network locations are shown in Figure S7, and were chosen in regions of parameter space that were likely to produce images that are not useful. As with the degraded images, logistic regression was used to determine the regions of usefulness likelihood on the contrast reduction vs. noise added parameter space. In the case of enhanced images, noise added was not a substantial predictor of classification (p_64×64_ = 0.09, p_128×128_ = 0.04), however contrast reduction was (p_64×64_ = 0.003, p_128×128_ = 8e-6). Despite this, the trend of the multivariate logistic regression for the enhanced images when compared to the same analysis of the degraded images offers some important insight.

As can be seen in Figure S13, the dashed lines indicate that the enhancement networks significantly increase the usefulness likelihood in degraded parameter space. For example, in the 128×128 parameter space, a portion of the line indicating degraded images that are 20% likely to be useful is predicted to be 90% likely to be useful after undergoing enhancement. The 90% line for 64×64 is less extreme, however the improvement from the network is still significant. The distance between lines of constant usefulness likelihood is also much larger for the enhancement networks, indicating slower drop-off toward the not useful region.

**Figure S13:**
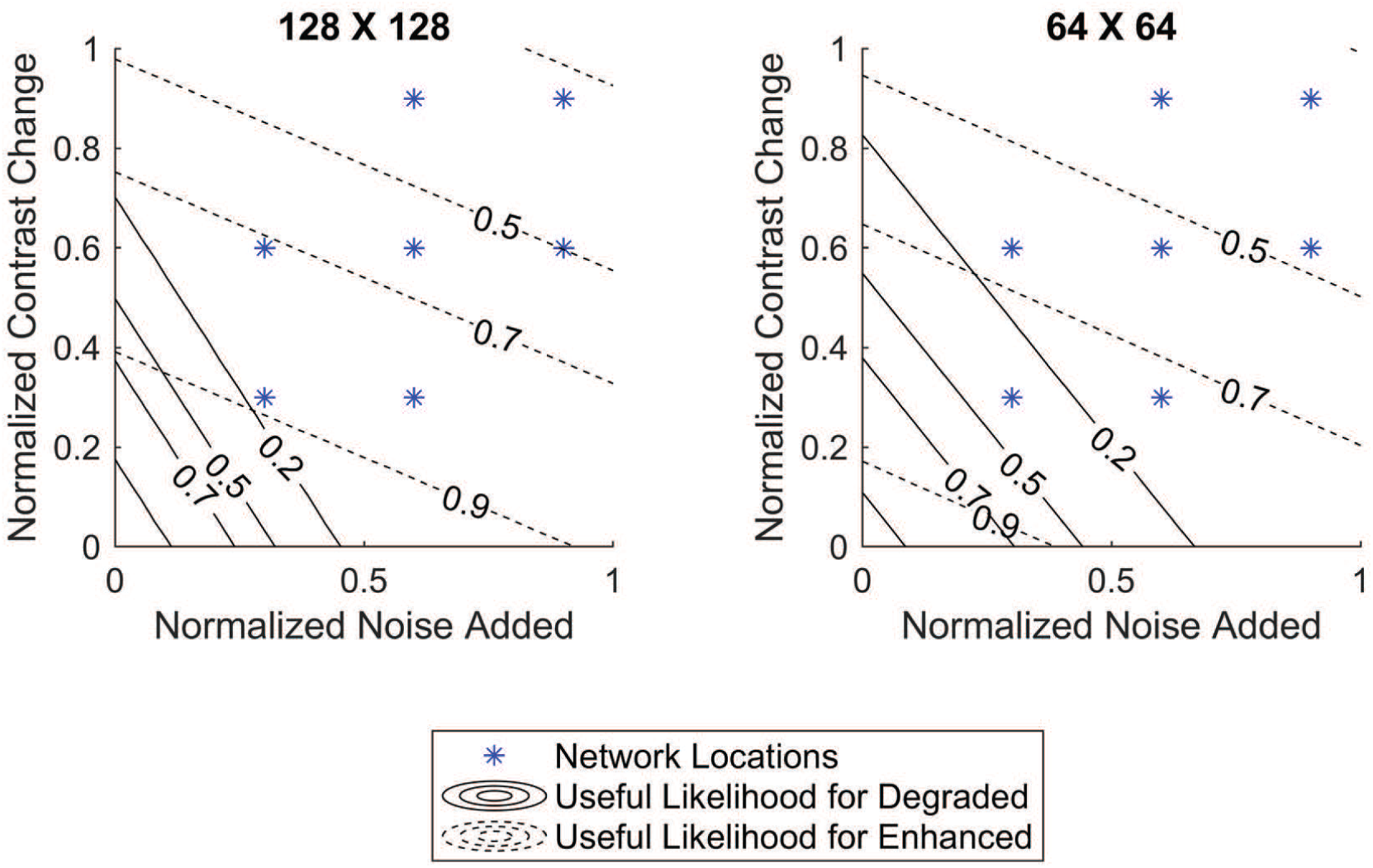
The combined classification of enhanced image data from the image utility assessment divided into 2 parameter planes of resolution 64×64 and 128×128. Solid lines of constant usefulness likelihood as determined by the logistic regression model for the degraded data are plotted with values indicated inline. Dashed lines of constant usefulness likelihood as determined by the logistic regression model for the enhanced data are also plotted. The enhancement networks appear to shift the usefulness likelihood into regions of greater degradation and rotate the slope with a preference toward noise added.

Another potentially important characteristic of Figure S13 is that the slope of the likelihood lines for the enhancement networks does not match the slope of the likelihood lines for the degraded images. The network seems to be skewed toward improving higher noise over higher contrast reduction. This could imply that the reconstruction errors could be a product of the networks inability to handle loss in contrast between brain and CSF. A network architecture more robust to addressing loss in contrast might yield fewer reconstruction errors. This concept represents a potential focus of research effort for the machine learning community to pursue in its application to biomedical imaging.

Since noise added was not significant for enhanced images, univariate logistic regression models were constructed for enhanced image classification with brain and CSF contrast prior to enhancement as the only predictor. Figure S14 shows the logistic regression models for 64×64 and 128×128 (p_64×64_ = 1e-6, p_128×128_ = 4e-9).

**Figure S14:**
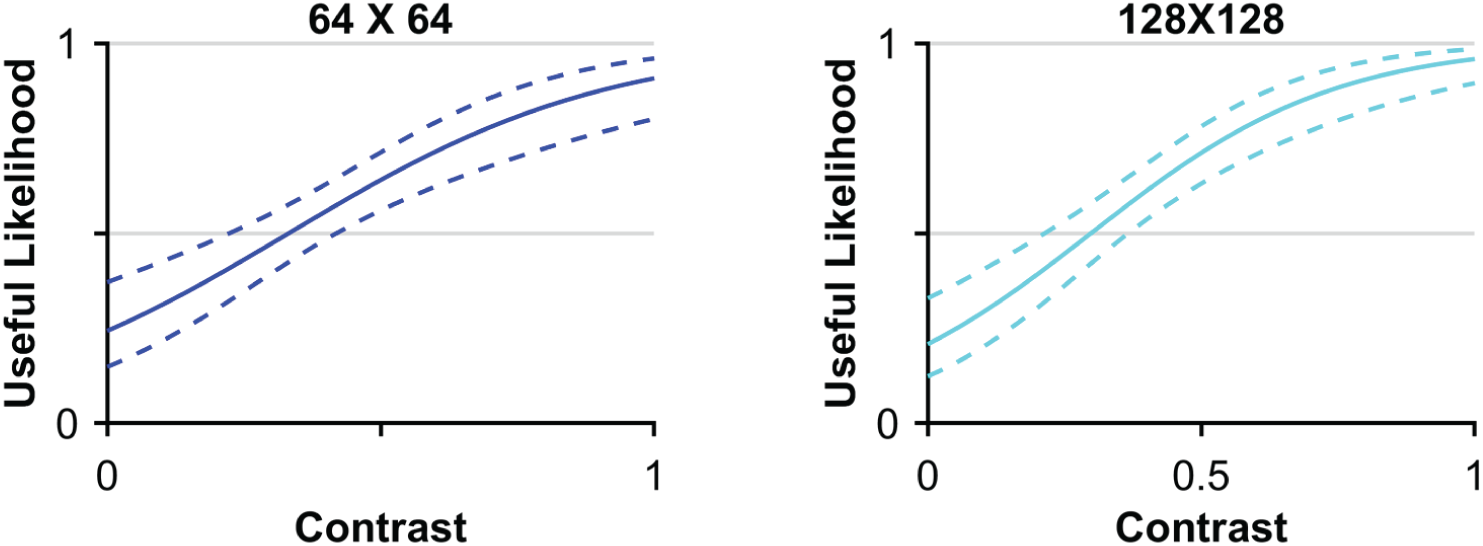
Logistic regression models of image utility with contrast between brain and CSF prior to enhancement as the single predictor for 64×64 and 128×128 enhanced images. The horizontal axis shows normalized contrast between brain and CSF with 0 being no contrast and 1 being the contrast found in the native CT images after downsampling via bilinear interpolation to 64×64 or 128×128 image resolution.

